# Differences in estimates for ten-year risk of cardiovascular disease in Black versus white persons with identical risk factor profiles using pooled cohort equations

**DOI:** 10.1101/2021.04.18.21255704

**Authors:** Ramachandran S. Vasan, Edwin van den Heuvel

## Abstract

**Background:** Sex- and race-specific pooled cohort equations (PCE) are recommended for estimating the 10-year risk of cardiovascular disease (CVD), with an absolute risk >7.5% indicating a clinical decision threshold.

**Methods:** We generated *in silico* 30,565 risk profiles in men and 29,515 in women by combining numerical (age, total and high-density lipoprotein cholesterol, systolic blood pressure) and binary risk factors (smoking, diabetes, antihypertensive treatment). We compared PCE-estimated 10-year CVD risk in Black versus white individuals with identical risk profiles. We performed similar comparisons in participants in the Framingham Third Generation cohort and the National Health and Nutrition Examination Survey 2017-2018.

**Results:** There were 6357 risk profiles associated with 10-year CVD risk >7.5% for Black but not for white men (median risk difference [RD] 6.25%, range 0.15-22.8%; median relative risk [RR] 2.40, range 1.02-12.6). There were 391 profiles with 10-year CVD risk >7.5% for white but not for Black men (median RD 2.68%, range 0.07-16.9%; median RR 1.42, range 1.01-3.57). There were 6543 risk profiles associated with 10-year estimated CVD risk >7.5% for Black but not for white women (median RD 6.14%, range 0.35-26.8%; median RR 2.29, range 1.05-12.6). There were 318 profiles with 10-year CVD risk >7.5% for white but not for Black women (median RD 3.71%, range 0.22-20.1%; median RR 1.66, range 1.03-5.46). The population-based samples demonstrated similar risk differences.

**Conclusions:** The PCE may generate substantially divergent CVD risk estimates for Black versus white individuals with identical risk profiles, which could introduce race-related variations in clinical recommendations for CVD prevention.

## Introduction

In 2013, the American College of Cardiology (ACC) and the American Heart Association (AHA) formulated sex- and race-specific equations (referred to as pooled cohort equations, PCE) for estimating the 10-year risk of atherosclerotic cardiovascular disease (CVD) using representative community-based cohorts.^1^ The PCE incorporate race and standard vascular risk factors, i.e., sex, age, systolic blood pressure, antihypertensive treatment, blood total cholesterol and high-density lipoprotein cholesterol concentrations, diabetes mellitus, and smoking. In 2019, the ACC/AHA endorsed the use of PCE in primary care settings to guide clinical decisions such as the prescription of statins for CVD prevention.^2^

In parallel, an emerging body of literature^3-10^ has criticized strongly the use of race in medical risk assessment because of concerns regarding algorithmic fairness,^11^ including the risk of exacerbating health inequities. Others have emphasized that prediction equations intended specifically to guide treatment decisions must incorporate causal risk factors.^12-14^ In this context, predictor variables in the PCE are biological in nature with the exception of race, which is a social construct.^3^ To our knowledge, a systematic analysis of the potential for race-related differential clinical decisions resulting from the use of PCE (for CVD risk assessments) has not been conducted hitherto. We evaluated this premise by comparing Black-white differences in PCE-estimated absolute CVD risk across a wide range of plausible risk factor combinations using an *in silico* approach that was complemented by observations in two community-based samples.

## Methods

We used hypothetical data for *in silico* analyses and de-identified public use data from the National Health and Nutrition Examination Survey (NHANES) 2017-2018. We used data from the Framingham Heart Study (FHS) Third Generation cohort participants at their first examination cycle. All FHS participants provided written informed consent, and the study protocol was approved by the Institutional Review Board at the Boston University Medical Center.

### Creating risk factor categories and their combinations

We considered a wide range of permissible values for CVD risk factors as specified for the ACC web-based risk estimator.^15^ We created profiles by combining CVD risk factors thus: age, 40 to ≤80 years, with 5 year increments (8 categories); systolic blood pressure, 100 to 200 mm Hg, with 10 mm Hg increments (10 categories); Total cholesterol concentrations, 130 to 290 mg/dL with 20 mg/dL increments (8 categories); high-density lipoprotein cholesterol (HDL) concentrations, 20 to 90 mg/dL, with 10 mg/dL increments (7 categories); diabetes mellitus (yes versus no; 2 categories); smoking status (yes versus no; 2 categories); treatment for elevated systolic blood pressure (yes versus no; 2 categories). We limited values of treated systolic blood pressure empirically to a maximum of 180 mm Hg. Thus, for each of four strata (men versus women; Black versus white persons), we created 51,840 possible risk factor combinations (also referred to as risk profiles).

### Estimating 10-year CVD risk for risk factor combinations with PCE

We calculated the 10-year absolute CVD risk for the 51,840 risk factor combinations using published PCE risk functions.^1^ We excluded risk profiles that yielded 10-year CVD risk estimates that were below 1% or above 30%, as recommended by the 2013 ACC/AHA guidelines.^1^

### Differences in PCE-estimated 10-year CVD risk for Black vs. white persons with the identical risk factor combinations

All analyses were sex-specific. First, we calculated differences in the PCE-based 10-year absolute risk and the relative risk of a CVD event for Black versus white persons with identical risk factor combinations. Next we evaluated *four possible scenarios* (two for each sex) in which the 10-year absolute CVD risk estimates for the two races diverged, being on opposite sides of the critical 7.5% 10-year CVD risk threshold that triggers clinical decisions (when exceeded), i.e., discussions with patients regarding initiating treatment with statins.^1,2^

For each scenario, we plotted histograms to describe the sex-specific distributions of the differences in absolute and relative risks (of CVD) for Black versus white people. We identified specific risk factor combinations that yielded divergent 10-year CVD risk estimates for Black versus white persons, including those that maximized these risk differences. We visualized the distributions of numerical risk factors (age, systolic blood pressure, total cholesterol and HDL cholesterol) for these combinations using sex-specific box plots. We also formulated sex-specific box plots showing 10-year absolute CVD risk differences for Black versus white persons according to the categories of each of the numerical risk factors.

Additionally, we repeated all our analyses using risk factor combinations within the ‘normal’ range, i.e., for people without diabetes mellitus, high blood pressure or history of smoking, and for values of systolic blood pressure 100-130 mm Hg, total cholesterol concentrations 130-170 mg/dl, and HDL concentrations 40-70 mg/dl in men or 50-80 mg/dl in women.

### Analysis of FHS data

We calculated the PCE-based 10-year CVD risk for 4086 white participants in the Third Generation FHS cohort^16^ using their risk factor data at their first examination cycle (2002-2005) and for their theoretical Black *counterfactuals* of the same sex (assuming they had an identical risk factor profile). We displayed the differences in absolute CVD risk for the four scenarios where white participants versus their Black counterfactuals had absolute CVD risk on opposite sides of the 7.5% risk threshold, paralleling our *in silico* approach.

### Analysis of NHANES 2017-2018 data

We accessed NHANES 2017-2018 public use data.^17^ We calculated the PCE-based 10-year CVD risk for Black and white participants with complete information available on their risk factor profiles, and for their same sex counterfactuals with identical risk profiles. We calculated displayed the differences in the PCE-estimated 10-year absolute CVD risk for these participants and for their counterfactuals where the factual and the counterfactual 10-year CVD risks were on opposite sides of the 7.5% risk threshold, consistent with our *in silico* approach.

## Results

### Risk factor combinations evaluated *in silico*

We evaluated 29515 and 30565 risk factor combinations for women and men, respectively (**Table S1**), after excluding profiles that generated 10-year CVD risk estimates below 1% or above 30%. In both sexes, 45% of the putative risk factor combinations included smoking whereas 40% included diabetes.

### Divergent PCE-estimated 10-year CVD risks in Black versus white men with identical risk factor profiles

There are 6357 risk factor combinations where a Black man has an estimated 10-year absolute CVD risk exceeding 7.5%, but a white man with a matched risk factor profile has an estimated CVD risk below that threshold (**Table S2**); the proportions of smoking and diabetes are 39.7% and 49.4%, respectively. **Figure 1 Panels A-B** present the histograms and summary statistics of Black-white differences in absolute and relative risk of CVD in men with these risk factor combinations. **Figure 1** demonstrates that the Black-white differences in absolute CVD risk can be as large as 22.8% (median 6.25%; **Panel A**), and the Black versus white relative risk for CVD can be as large as 12.6 (median 2.40; **Panel B**).

**Figure1, Panel A.**
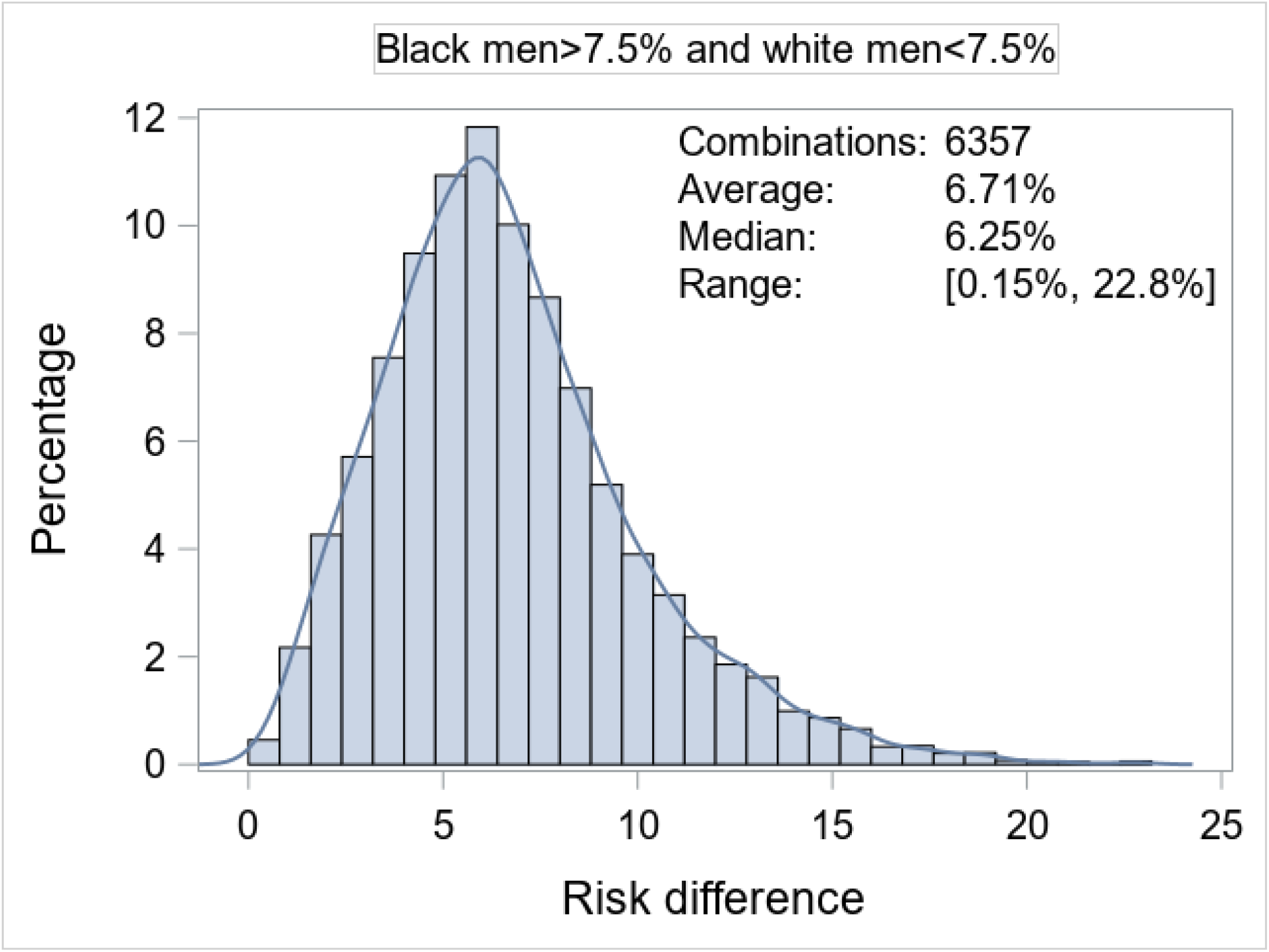
Distributions of Black-white differences in pooled cohort equation estimated 10-year CVD risk in men when absolute CVD risk exceeds 7.5% in Black but not in white men.

**Figure 1, Panel B.**
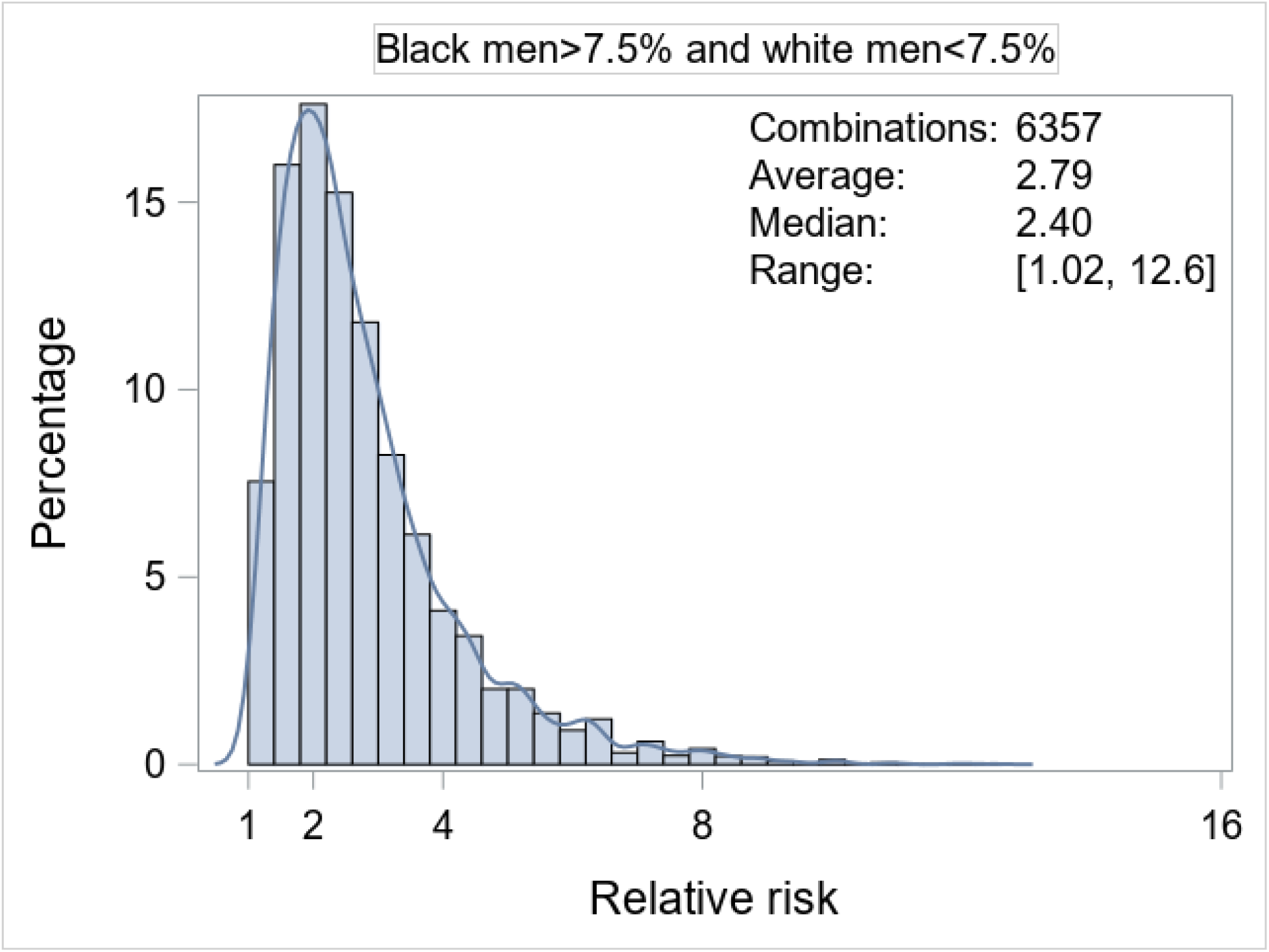
Distributions of relative risks (black versus white men) for 10-year incidence of CVD when pooled cohort equation estimated absolute risk exceeds 7.5% in Black but not in white men.

There are 391 risk factor combinations where a white man has an estimated absolute CVD risk exceeding 7.5% but a black man with an identical risk factor profile would have an estimated risk below that threshold (**Table S2)**; the proportions of smoking and diabetes are 32.5% and 7.2%, respectively. **Figure 1 Panels C-D** demonstrate that the white-Black differences in absolute CVD risk can be as large as 16.9% (median 2.68%) and the white versus Blacks relative risk of CVD can be as large as 3.57 (median 1.42).

**Figure 1, Panel C.**
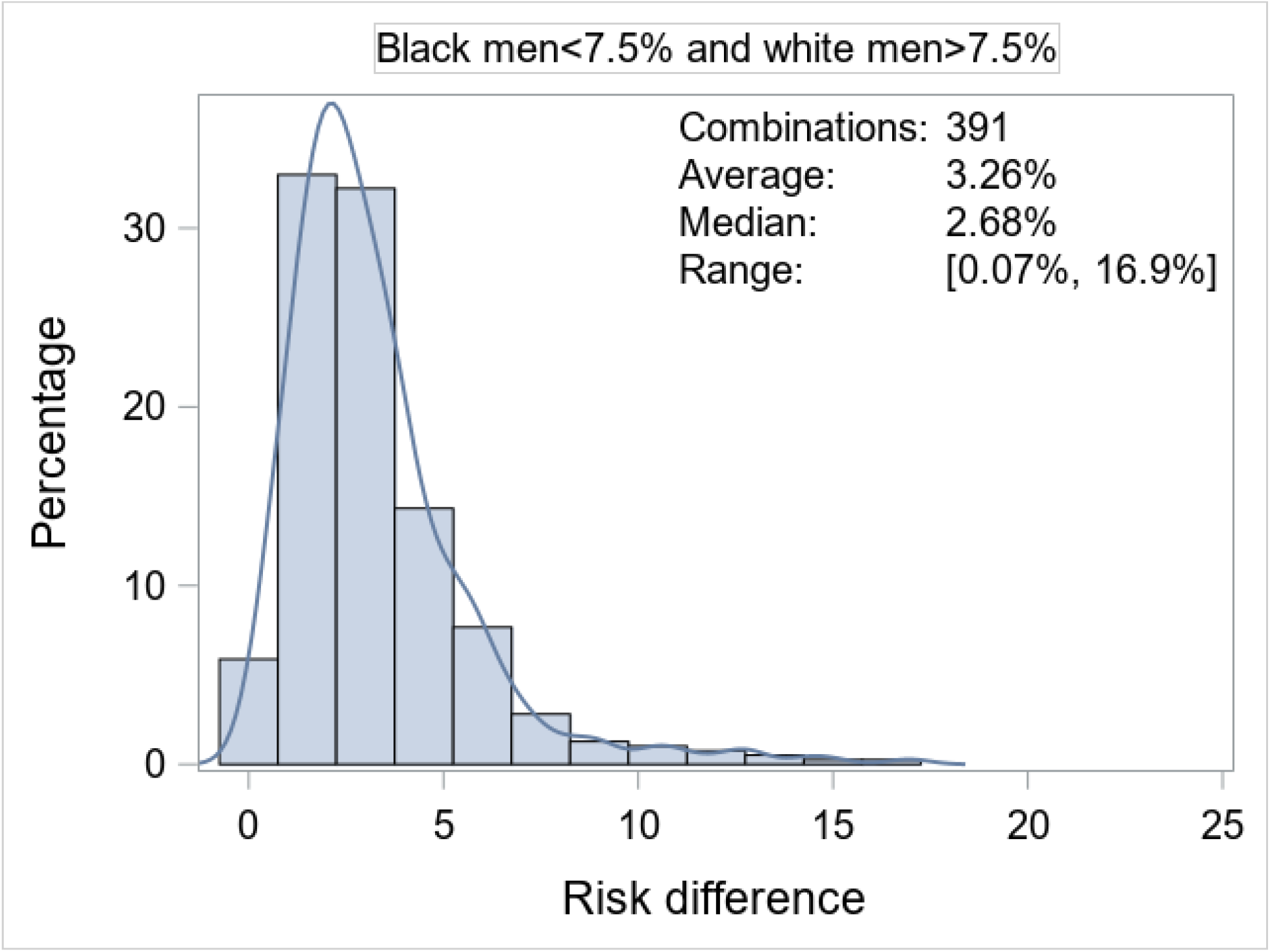
Distributions of white-Black differences in pooled cohort equation estimated 10-year CVD risk in men when absolute CVD risk exceeds 7.5% in white but not in black men.

**Figure 1, Panel D.**
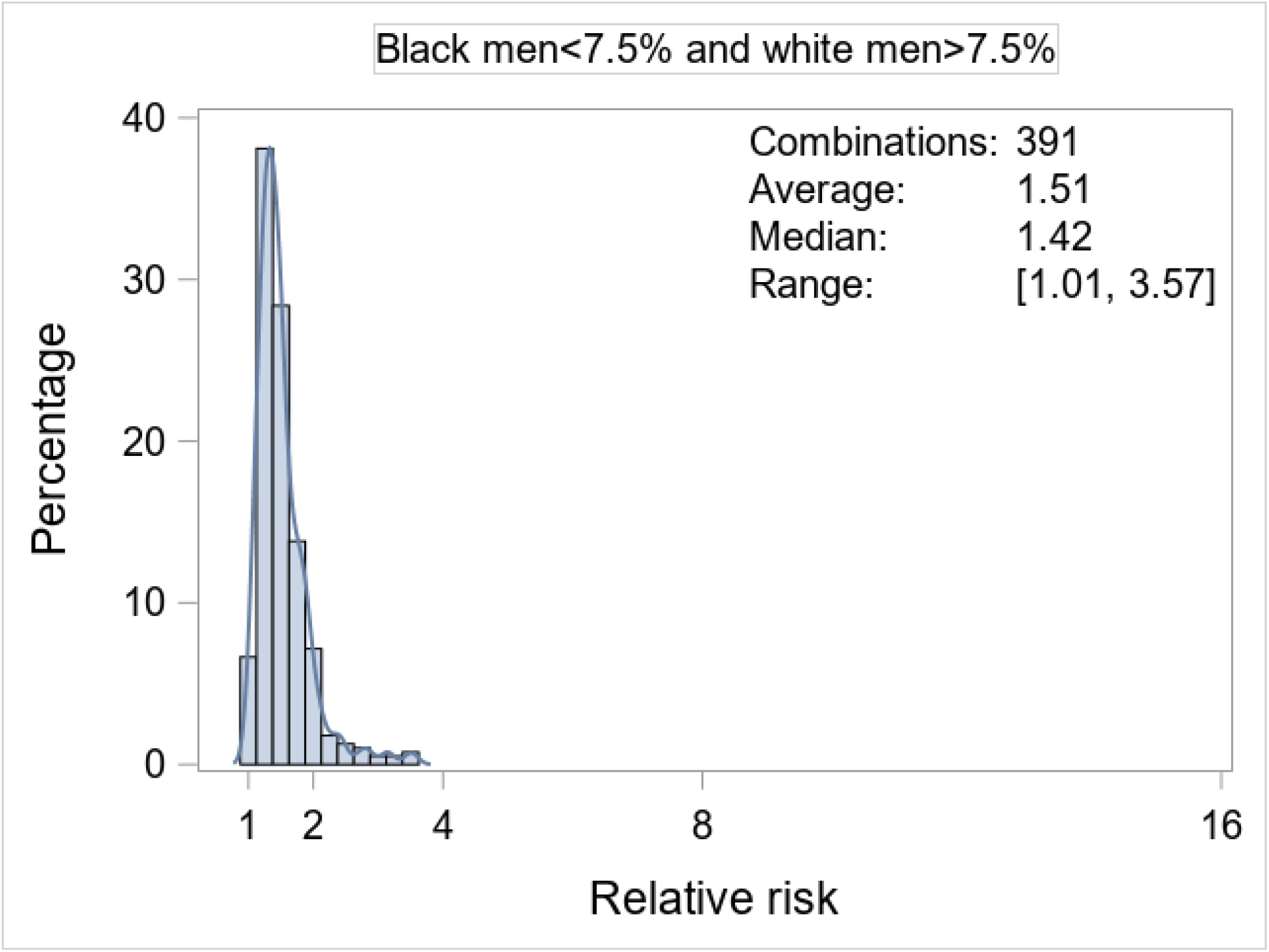
Distributions of relative risks (white versus Black men) for 10-year incidence of CVD when pooled cohort equation estimated absolute risk exceeds 7.5% in white but not in Black men.

**Table 1 (parts A-B)** shows specific risk factor combinations that yield maximal Black-white differences in absolute and relative risks of CVD in men (within the constraints of our inclusion criteria). **Figure S1** displays the box plot distributions of numerical risk factors for combinations that yield divergent (and convergent) CVD risk estimates for Black versus white men. **Figure S2** shows box plots for the Black-white differences in absolute CVD risk estimates for various risk factor categories. Risk factor combinations that yield an estimated 10-year CVD risk of 7.5% or more for Black but not for white men typically occur at younger ages, and with a higher frequency of non-smokers, and at higher values of systolic blood pressure. Risk factor combinations that yield an absolute CVD risk exceeding 7.5% for white but not for Black men are associated with a high prevalence of hypertension treatment, lower HDL levels, and absence of diabetes and smoking. Even with risk factor values within the normal range, there are profiles that can yield moderate-to-large Black-white differences in absolute CVD risk (**Table S3)**.

**Table 1.** Risk factor combinations for which the difference in 10-year pooled cohort equation estimated risk and relative risk of CVD are maximal for Black versus white individuals (or vice versa)

| 10-year CVD Risk Measure | Age, years | Hypertension | Smoking | Diabetes | Systolic BP, mm Hg | Total Cholesterol, mg/dl | HDL, mg/dl | 10-year CVD Risk % (PCE) |  | Maximal Value* |
| --- | --- | --- | --- | --- | --- | --- | --- | --- | --- | --- |
|  |  |  |  |  |  |  |  | Blacks | Whites |  |
| A. Black men are at risk<sup>a</sup> but white men are not |  |  |  |  |  |  |  |  |  |  |
| Black-White, Risk Difference | 45 | 1 | 1 | 1 | 180 | 130 | 80 | 28.8% | 6.00% | 22.8% |
| Black/White, Relative Risk | 40 | 1 | 0 | 1 | 180 | 150 | 90 | 13.8% | 1.10% | 12.6 |
| B. White men are at risk<sup>a</sup> but Black men are not |  |  |  |  |  |  |  |  |  |  |
| White-Black, Risk Difference | 40 | 0 | 1 | 0 | 120 | 290 | 20 | 6.92% | 23.9% | 16.9% |
| White/Black Relative Risk | 40 | 0 | 1 | 0 | 100 | 290 | 20 | 5.03% | 17.9% | 3.57 |
| C. Black women are at risk<sup>a</sup> but white women are not |  |  |  |  |  |  |  |  |  |  |
| Black-White, Risk Difference | 40 | 1 | 0 | 1 | 160 | 130 | 30 | 29.9% | 3.17% | 26.8% |
| Black/White, Relative Risk | 40 | 1 | 0 | 1 | 180 | 130 | 40 | 29.0% | 2.30% | 12.6 |
| D. White women are at risk<sup>a</sup> but Black women are not |  |  |  |  |  |  |  |  |  |  |
| White-Black, Risk Difference | 80 | 1 | 0 | 0 | 120 | 130 | 20 | 6.38% | 26.4% | 20.1% |
| White/Black, Relative Risk | 40 | 0 | 1 | 0 | 100 | 290 | 30 | 1.83% | 9.99% | 5.46 |

<sup>a</sup>>7.5% 10-year risk of atherosclerotic hard CVD as defined by the PCE (pooled cohort equations).
\*refers to maximal value for risk difference or relative risk (according to row).
BP= blood pressure. HDL= high-density lipoprotein cholesterol
For binary risk factors, 1= present, 0= absent.

### Divergent PCE-estimated 10-year CVD risks in Black versus white women with identical risk factor profiles

There are 6543 risk factor combinations associated with a 10-year absolute risk estimate greater than 7.5% in Black but not in white women with identical risk factor profiles (**Table S4**); the proportions of smoking and diabetes are 31.1% and 48.3%, respectively. **Figure 2 Panels A-B** present the histograms and summary statistics of the Black-white differences in 10-year absolute and relative risk of CVD for these risk factor combinations. It is evident that the Black-white differences in absolute CVD risk can be as large as 26.8% (median 6.14%; **Panel A**) and the Blacks versus white relative risk of CVD can be as large as 12.6 (median 2.29; **Panel B**). There are 318 risk factor combinations that are associated with estimated 10-year absolute CVD risk that exceeds 7.5% in white but not in Black women (**Table S4**); the proportions of smoking was 68.2% while that of diabetes was 38.7%. **Figure 2 Panels C-D** demonstrates that the white-Black differences in estimated absolute CVD risk can be as large as 20.1% (median 3.17%) and the white versus Black relative risks of CVD can be as large as 5.46 (median 1.66). **Table 1 (parts C-D)** shows select risk factor combinations that yield maximal Black-white differences in estimated absolute and relative risks of CVD in women (within the boundaries of our inclusion criteria). **Figure S3** displays the box plots for numerical risk factors for the combinations that yield divergent (and convergent) estimated absolute CVD risk for Black versus white women. **Figure S4** shows the box plots for the differences in PCE-estimated 10-year CVD risk for black versus white women for the individual risk factor categories. Estimated 10-year CVD risk exceeds 7.5% in Black but not white women more frequently in non-smoking adults and when diabetes is present. Conversely, a higher estimated absolute CVD risk for white but not Black women occurs more frequently among smokers. Even across the normal range of risk factors, we observe moderate differences in absolute CVD risk estimates for Black versus white women (**Table S5**).

**Figure 2, Panel A.**
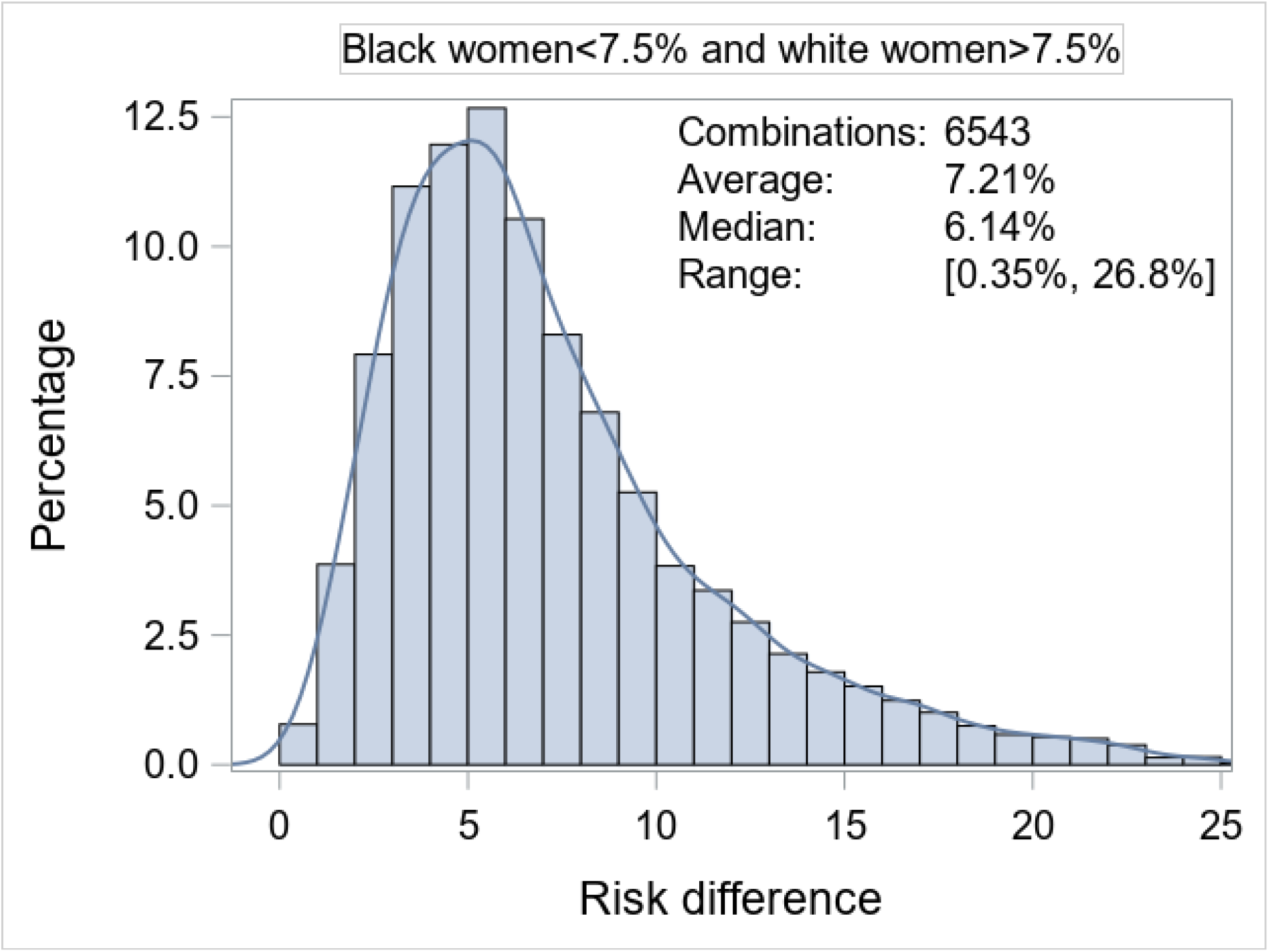
Distributions of Black-white differences in pooled cohort equation estimated 10-year CVD risk in women when absolute CVD risk exceeds 7.5% in Black but not in white women.

**Figure 2, Panel B.**
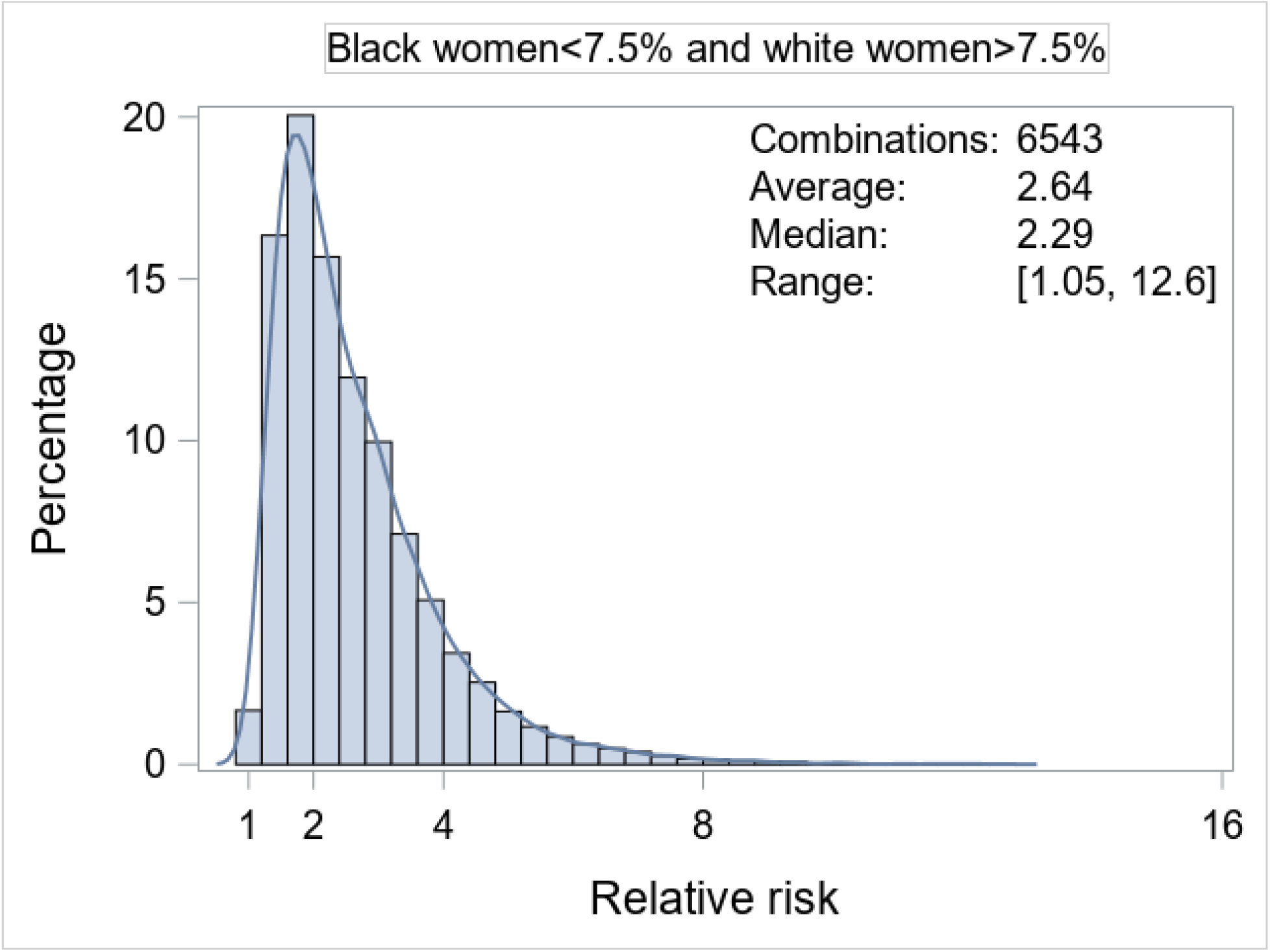
Distributions of relative risks (black versus white women) for 10-year incidence of CVD when pooled cohort equation estimated absolute risk exceeds 7.5% in Black but not in white women.

**Figure 2, Panel C.**
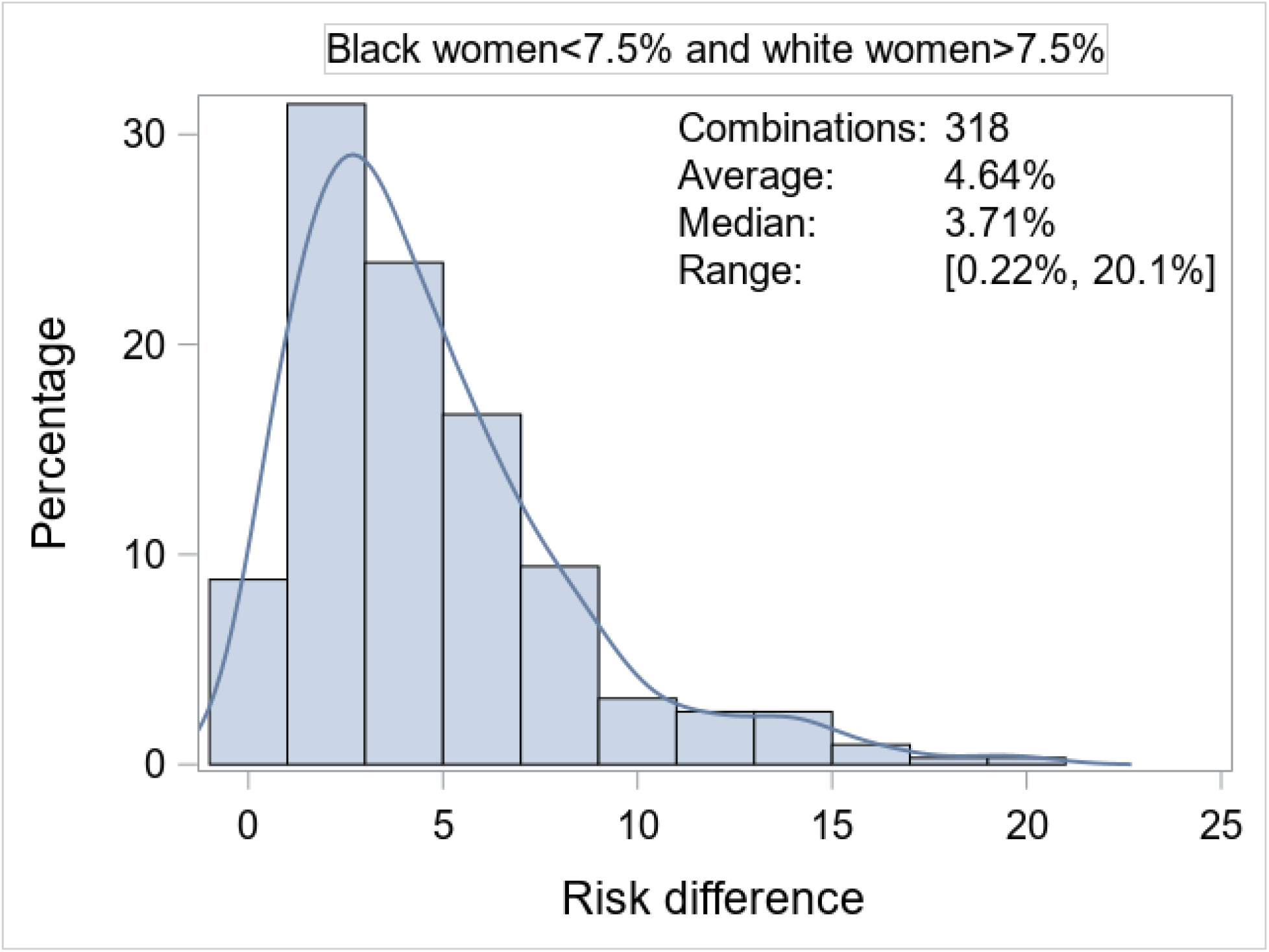
Distributions of white-Black differences in pooled cohort equation estimated 10-year CVD risk in women when absolute CVD risk exceeds 7.5% in white but not in black women.

**Figure 2, Panel D.**
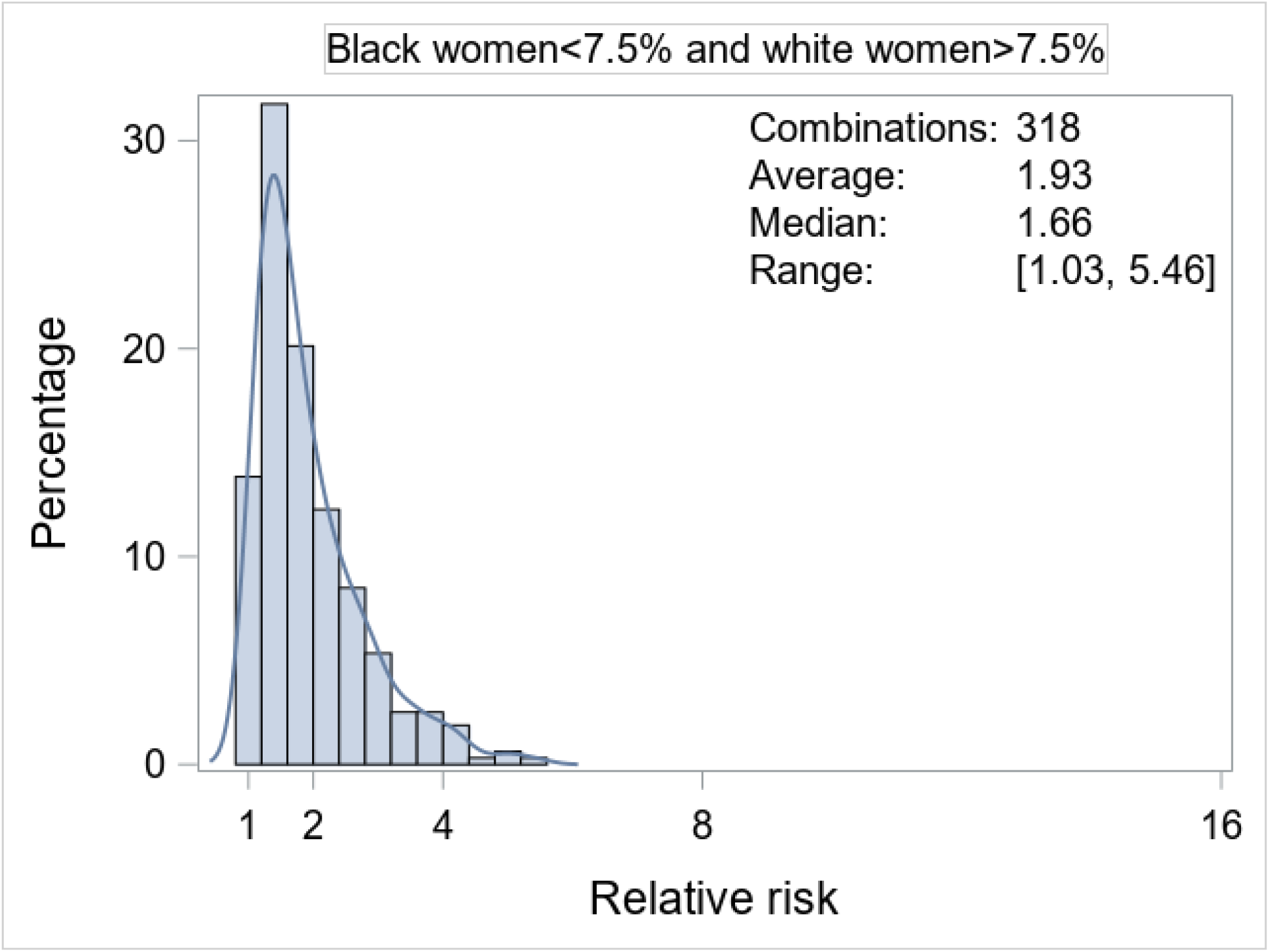
Distributions of relative risks (white versus Black women) for 10-year incidence of CVD when pooled cohort equation estimated absolute risk exceeds 7.5% in white but not in Black women.

### PCE-based 10-year CVD risk estimates for white FHS participants and their Black counterfactuals

Of 4086 white FHS Third Generation participants (**Table S6**), we excluded 1901 individuals below age 40 years, 258 participants with risk factor values outside the range for our *in silico* analysis, and 619 people with PCE-based risk estimates outside the recommended range per guidelines.^1^ After exclusions, 1273 participants (378 women) remained eligible for calculation of PCE-based CVD risk estimates. Of these, 121 white participants (9.5%; 34 women) had an estimated 10-year CVD risk below 7.5%, while their Black counterfactuals had an estimated absolute CVD risk above that threshold. Conversely, 17 white individuals (1.34%; 2 women) had an estimated 10-year CVD risk above 7.5%, while their Black counterfactuals had an estimated absolute risk below that threshold. **Figure 3** shows the distributions of estimated absolute CVD risk differences between white FHS participants and their Black counterfactuals.

**Figure 3, Panel A.**
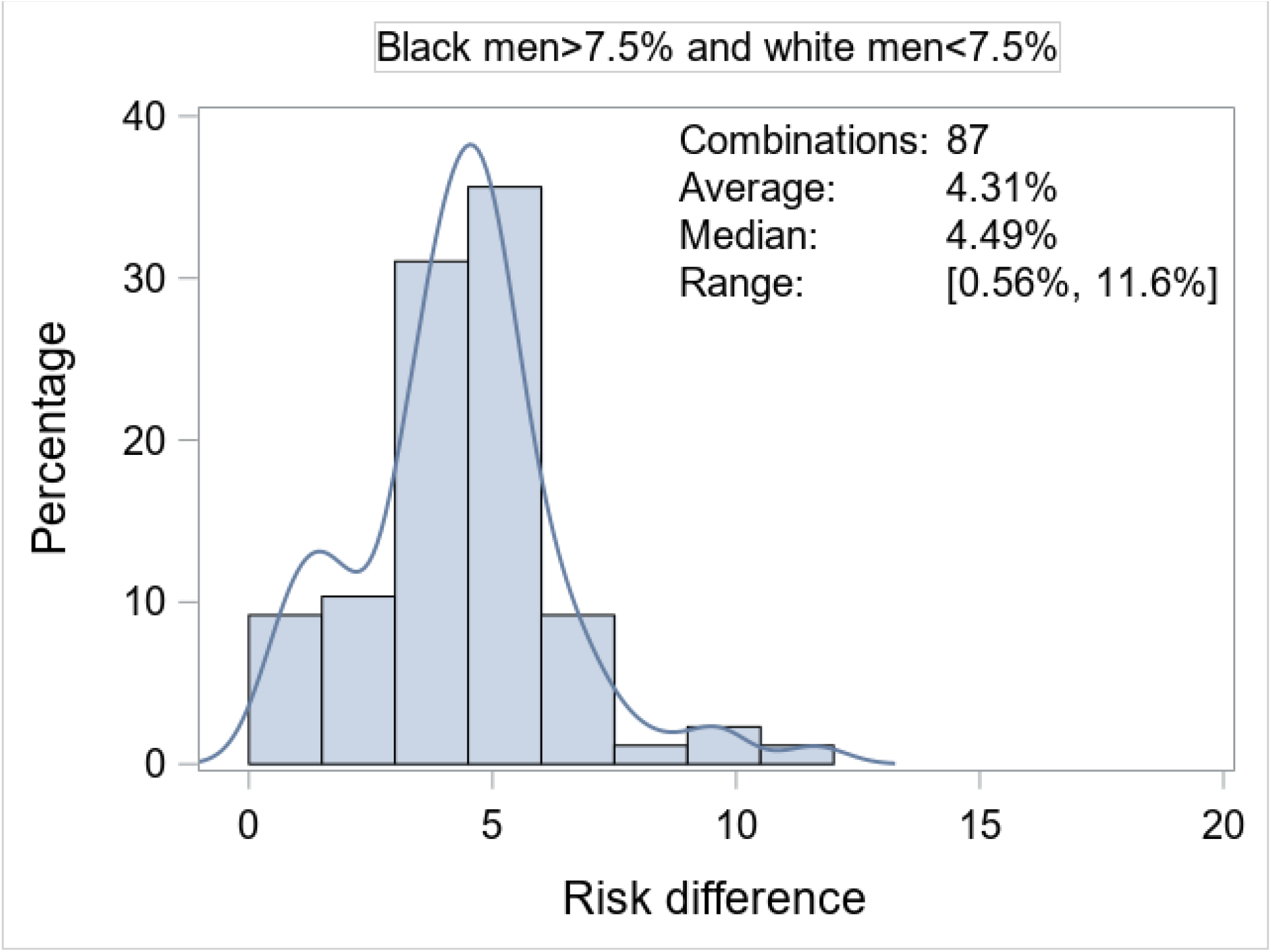
Distribution of Black-white differences in pooled cohort equation estimated 10-year CVD risk when absolute CVD risk does not exceed 7.5% in white men in the Framingham Heart Study but exceeds this threshold in their Black counterfactuals.

**Figure 3, Panel B.**
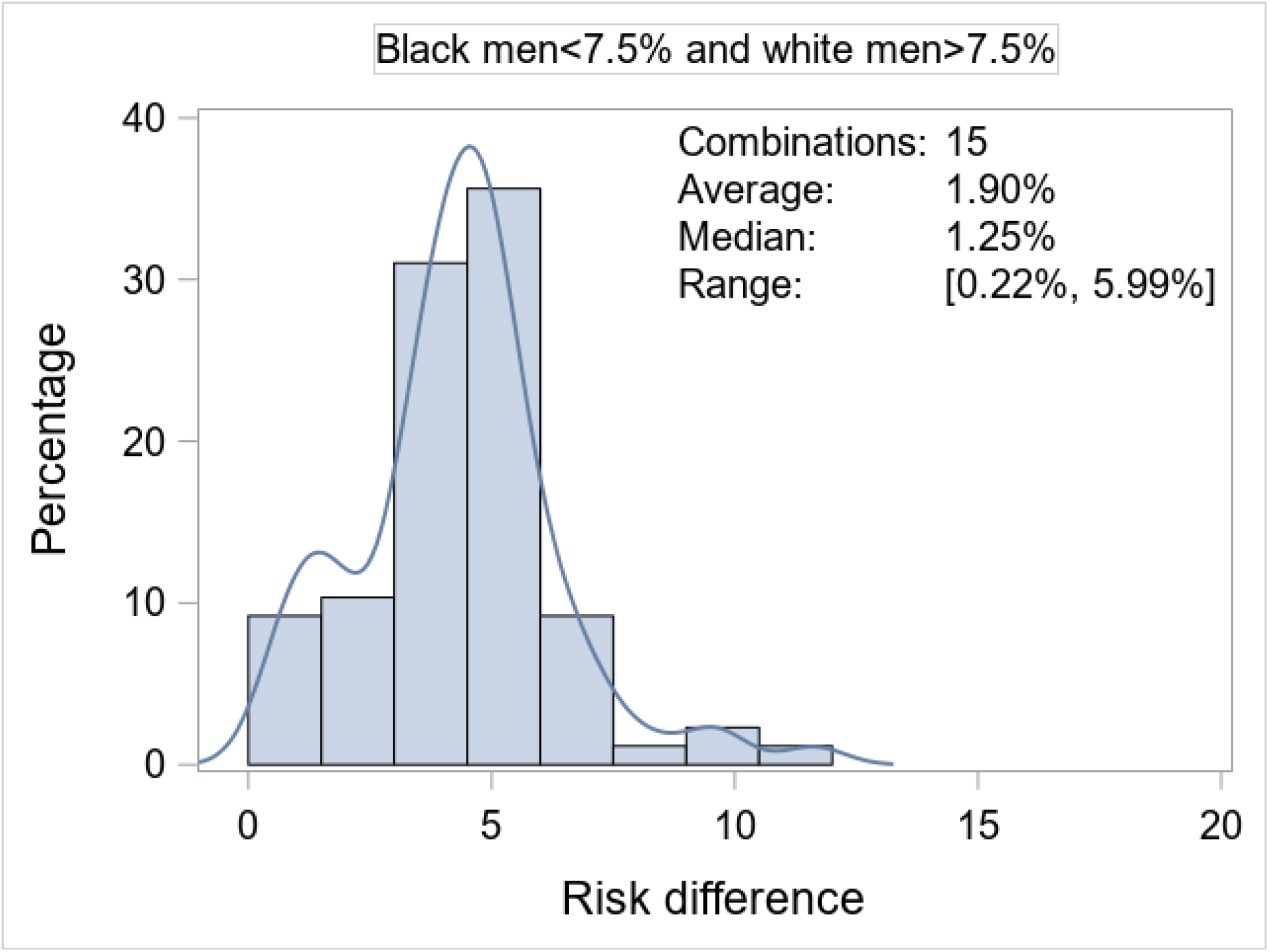
Distribution of white-Black differences in pooled cohort equation estimated 10-year CVD risk when absolute CVD risk exceeds 7.5% in white men in the Framingham Heart Study but does not exceed this threshold in their Black counterfactuals.

**Figure 3, Panel C.**
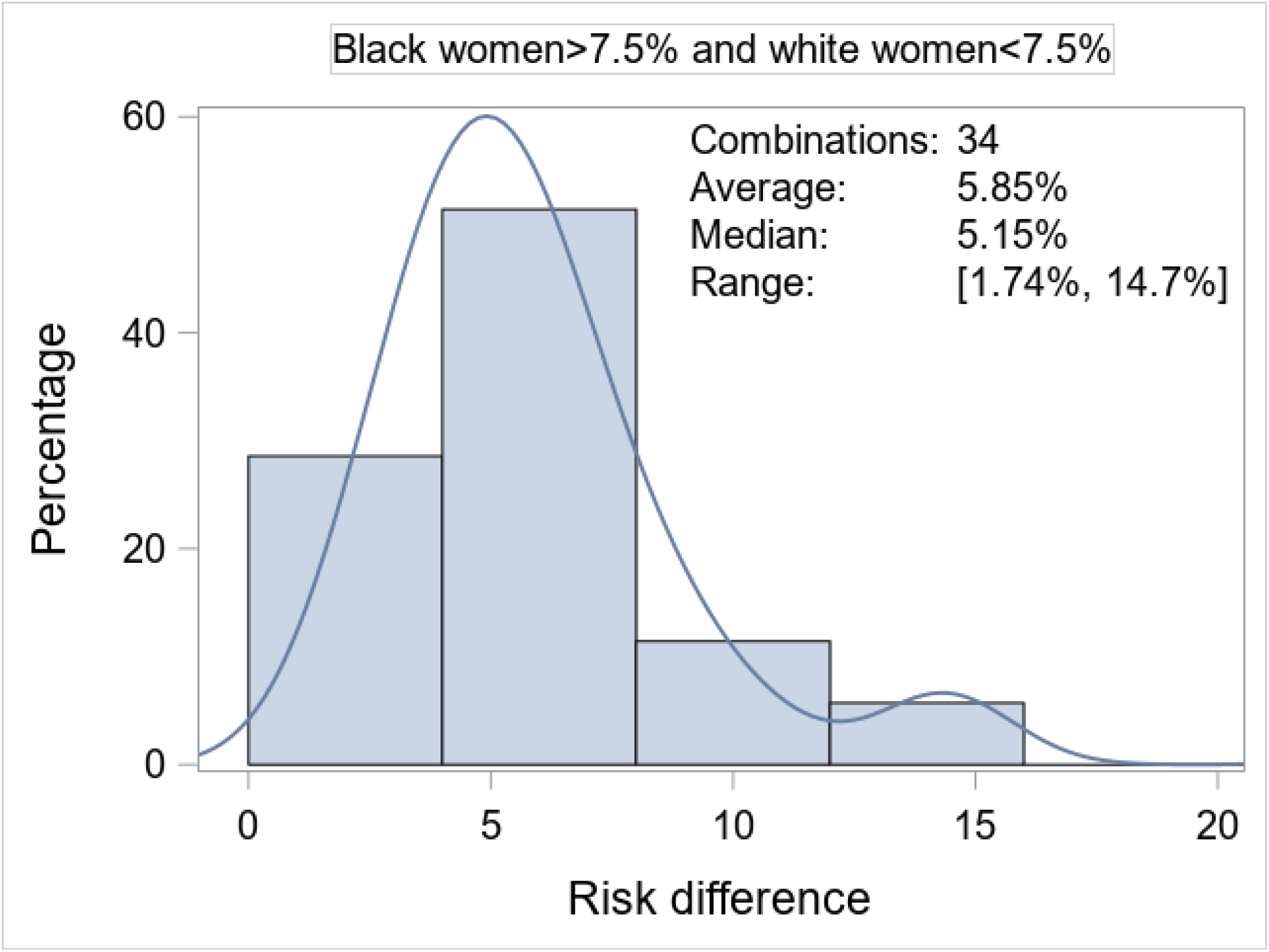
Distribution of Black-white differences in pooled cohort equation estimated 10-year CVD risk when absolute CVD risk does not exceed 7.5% in white women in the Framingham Heart Study but exceeds this threshold in their Black counterfactuals.

**Figure 3, Panel D.**
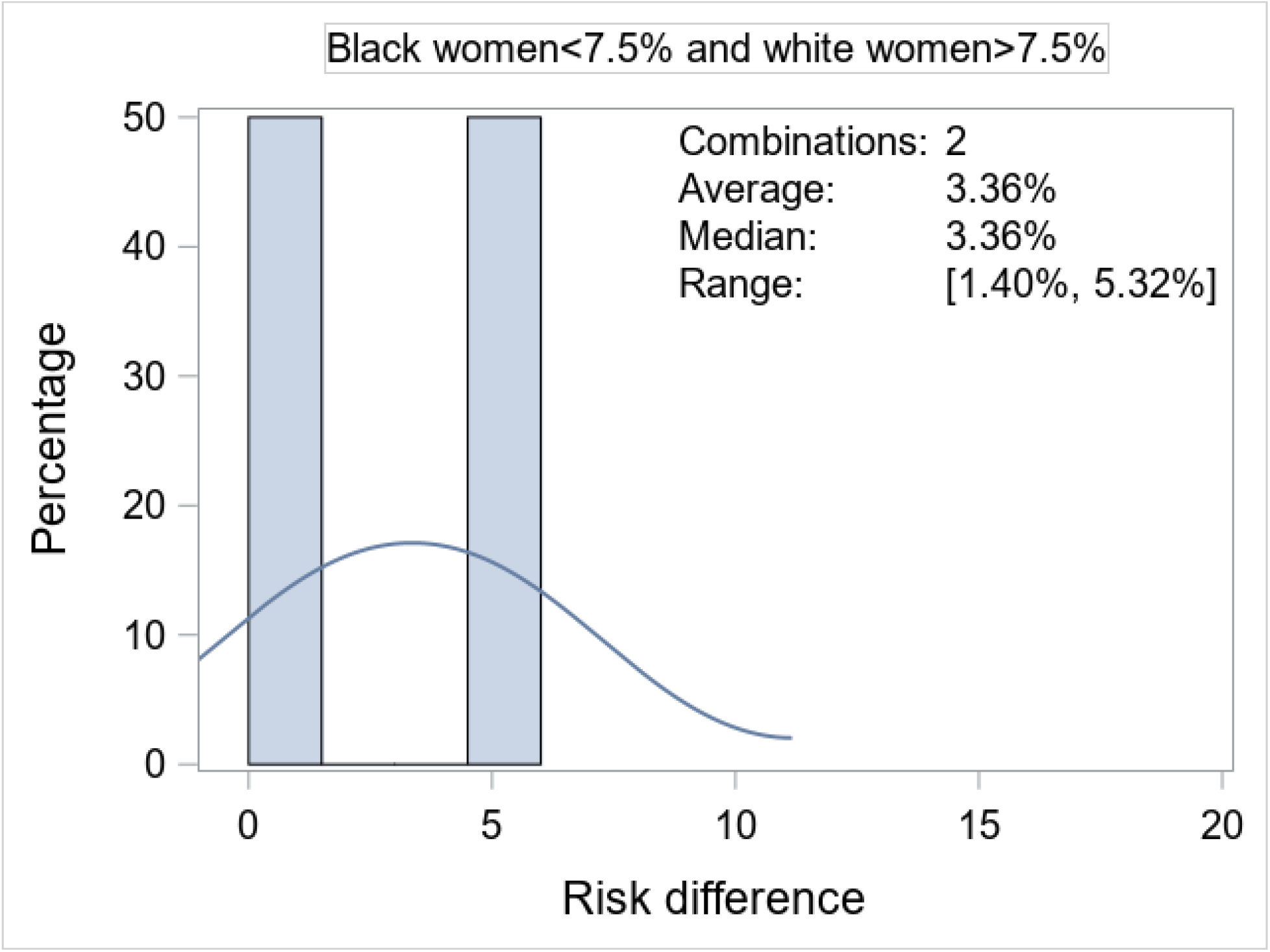
Distribution of white-Black differences in in pooled cohort equation estimated 10-year CVD risk when absolute CVD risk exceeds 7.5% in white women in the Framingham Heart Study but does not exceed in their Black counterfactuals.

### PCE-based 10-year CVD risk estimates for NHANES participants and their counterfactuals

Of 1065 NHANES participants (**Table S7**), we excluded 365 individuals with risk factor values outside the range for our *in silico* analysis, and 150 people with PCE-based risk estimates outside the recommended range per guidelines.^1^ After exclusions, 550 participants (223 women, 36.8% Blacks; and 327 men, 40.4% Blacks) remained eligible for calculation of PCE-based CVD risk estimates. Of these participants, 34 white participants (10.1%; 19 women) had an estimated 10-year CVD risk below 7.5%, while their Black counterfactuals had an estimated absolute CVD risk above that threshold (**Table 2**). Conversely, seven white individuals (2.1%; 1 women) had an estimated 10-year CVD risk above 7.5%, while their Black counterfactuals had an estimated absolute risk below that threshold (**Table 2**). Furthermore, 31 Black participants (14.5%, 19 women) had an estimated CVD risk above 7.5% while their white counterfactuals had an estimated CVD risk below 7.5% (**Table 2**). Only one Black participant (1 women) had an estimated risk below 7.5% with a counterfactual white risk of CVD above 7.5% (**Table 2**).

**Table 2.**
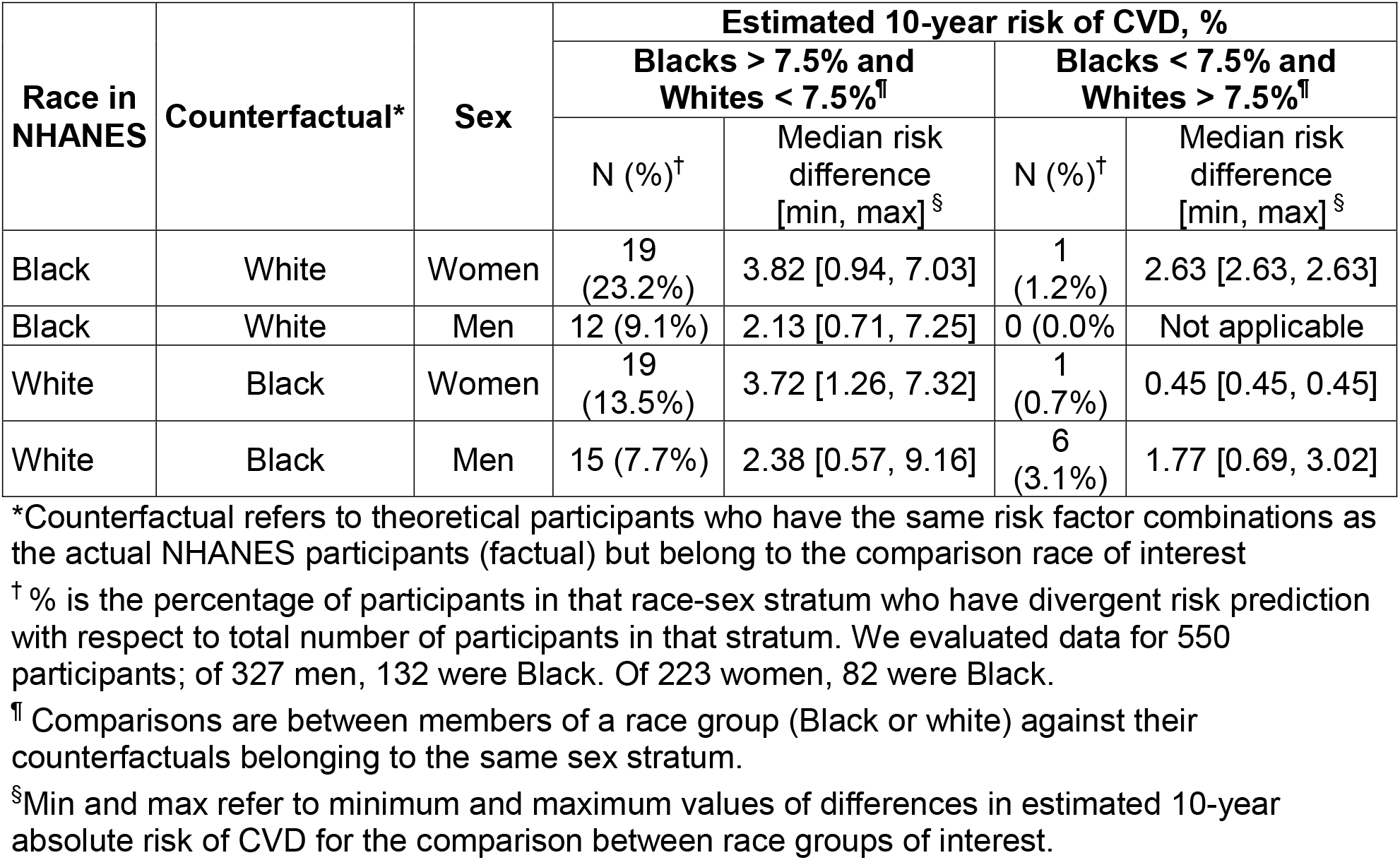
Summary statistics for divergent pooled cohort equations-based estimated 10-year CVD risk in white and black participants of the NHANES 2017-2018 cycle.

| Race in NHANES | Counterfactual* | Sex | Estimated 10-year risk of CVD, % |  |  |  |
| --- | --- | --- | --- | --- | --- | --- |
|  |  |  | Blacks > 7.5% and Whites < 7.5%<sup>¶</sup> |  | Blacks < 7.5% and Whites > 7.5%<sup>¶</sup> |  |
|  |  |  | N (%)<sup>†</sup> | Median risk difference [min, max]<sup>§</sup> | N (%)<sup>†</sup> | Median risk difference [min, max]<sup>§</sup> |
| Black | White | Women | 19 (23.2%) | 3.82 [0.94, 7.03] | 1 (1.2%) | 2.63 [2.63, 2.63] |
| Black | White | Men | 12 (9.1%) | 2.13 [0.71, 7.25] | 0 (0.0%) | Not applicable |
| White | Black | Women | 19 (13.5%) | 3.72 [1.26, 7.32] | 1 (0.7%) | 0.45 [0.45, 0.45] |
| White | Black | Men | 15 (7.7%) | 2.38 [0.57, 9.16] | 6 (3.1%) | 1.77 [0.69, 3.02] |

\*Counterfactual refers to theoretical participants who have the same risk factor combinations as the actual NHANES participants (factual) but belong to the comparison race of interest
<sup>†</sup> % is the percentage of participants in that race-sex stratum who have divergent risk prediction with respect to total number of participants in that stratum. We evaluated data for 550 participants; of 327 men, 132 were Black. Of 223 women, 82 were Black.
<sup>¶</sup> Comparisons are between members of a race group (Black or white) against their counterfactuals belonging to the same sex stratum.
<sup>§</sup>Min and max refer to minimum and maximum values of differences in estimated 10-year absolute risk of CVD for the comparison between race groups of interest.

## Discussion

### Principal Findings

Our main findings are three-fold. First, use of PCE can result in major differences in predicted CVD risk for Black versus white people who have identical risk factor profiles, a situation that may occur frequently, as observed both *in silico* and in two independent community-based samples. In both sexes, PCE-estimated CVD risk estimates for Black persons exceeded that for white individuals with the same risk profile more frequently than the converse. The magnitude of these Black versus white risk differences (median 6%) and relative risks (median 2.3) are substantial, and seem biologically implausible.

Second, specific risk factor combinations in each sex may exacerbate Black-white differences in PCE-estimated absolute CVD risks. In both sexes, higher PCE-based CVD risk estimates for Black persons (relative to white individuals with identical risk factor profiles) were evident in younger age groups, with higher values of systolic blood pressure, and when diabetes was present. In the older age groups and in the presence of low HDL concentrations, PCE-estimated CVD risk was higher in white compared to Black women with identical risk factor profiles. Risk factor combinations with lower total cholesterol or higher HDL concentrations were associated with higher PCE-estimated CVD risks in Black versus white men.

Third, such race-related differences in PCE-estimated CVD risk may be clinically meaningful― they could result in different treatment decisions (such as statin prescription) in people with identical risk factor profiles based solely on their race.

### Comparison with the published literature

It is widely acknowledged that PCE can both under- and over-estimate CVD risk in different contexts.^18-25^ Yet data are scant regarding how frequently PCE generates considerably divergent risk estimates for Black versus white persons *with identical risk factor profiles that could impact clinical decisions*. Yadlowski et al.^25^ reported divergent PCE-derived CVD risk estimates for Black versus white persons in the NHANES 2013-2014 sample. The authors^25^ noted that updating the PCE with data from recent cohort samples, and reducing model overfitting attenuated but did not eliminate these race-related differences.^25^ To our knowledge, no prior report has evaluated whether specific risk factor combinations may exaggerate Black-white differences in PCE-estimated CVD risk.

### Clinical Implications

First, the use of PCE may result in Black persons with select risk factor combinations becoming more eligible for receiving statin treatment compared to their white counterparts with identical risk profiles. Although the directionality of this potential bias may be somewhat reassuring (relative to the opposite scenario of Black persons not receiving statins relative to their white counterparts), the risks associated with overtreatment (financial, psychological, side effects, quality of life, etc.) may not be trivial.

Second, race is a social construct created by humans “to group people with certain observable physical characteristics, such as skin color or facial features, who evolved from different geographies in the world.”^3^ It is widely accepted also that race is associated with health outcomes and health inequities, in part mediated by “exposure or vulnerability to behavioral, psychosocial, material, or environmental risk factors and resources.”^26^ Thus, Black-white differences in PCE-based CVD risk estimates may be a surrogate for structural racism, differences in health care access, educational achievement and economic challenges, and other sources of health inequities.^27^ Therefore, race should be replaced in any risk prediction equation by the assortment of potentially causal factors it represents and that can be intervened upon. If replaced by appropriate causal variables, race should no longer improve prediction in the risk algorithms. For instance, at least three UK-based risk scores^28-30^ incorporate a ‘social deprivation index’ (rather than race) that more directly addresses the social determinants of health.

Third, it is critical to make a distinction between risk prediction unrelated to interventional decisions (e.g. for prognostication) and *prediction equations that are tied to decisions for intervention* (such as the PCE).^13^ The latter situation requires a causal framework for multiple reasons,^14^ including the importance of addressing true *root causes*, enhancing *transportability* of the prediction algorithm across settings^12^ and ensuring *prediction invariance*.^31^ The latter refers to the concept that a prediction from a causal model will work as well under interventions as it does for observational data.^31^

Fourth, some scientists^32^ have argued that from an outcomes perspective, the incorporation of race in risk estimation algorithms may permit better concordance with the patient’s own goals, thereby facilitating individualized and optimal care. On the other hand, causal definitions of fairness would argue that comparable individuals (i.e., with identical risk factor profiles) should not be treated differentially based on attributes that may predispose them to discrimination, such as a certain race/ethnicity. Thus, it is imperative to generate expert consensus regarding how a clinician should optimally balance an algorithmic fairness framework against an outcomes-based approach.

Scientists have stipulated several criteria for including race in clinical prediction algorithms: the race-based measures are ‘reproducible at an individual level’; there is a sound scientific rationale for a causal role of race in disease etiology that is supported by robust scientific and statistical evidence; there are data indicating how such incorporation (of race) would mitigate rather than exacerbate harm for a group that is at greater risk of poor health outcomes; there is evidence that such benefit cannot be achieved ‘by other feasible means’; it is transparent, and accommodates in a fair manner individuals who ‘reject race categorization’.^4,5,10^

Fifth, additional research is needed also to evaluate if refinements to the PCE are necessary in select scenarios (e.g., for specific risk factor combinations) that are associated with race-related differences in risk estimates that are extreme and deemed to be biologically implausible. For instance, risk calculators could offer either *race-less* predictions (using variables other than race, ‘leaving the clinician to make their best judgment’)^*3*^ or they could provide estimates of predicted risk for both Black and white persons, regardless of the race of the person in front of the clinician. In the latter situation, the risk estimates for people of both races may be divergent but below a threshold when no clinical intervention is suggested (e.g., <5% 10-year CVD risk in both whites and Blacks) or above a level that requires action on the part of the clinician (e.g., >10% 10-year CVD risk in both whites and Blacks). In these circumstances, the race-related differences may result in ‘non-polar’ decisions, rendering the variances somewhat less germane. However, there may be zones of 10-year CVD risk estimates (e.g., people at intermediate risk with a 5%-10% PCE-based absolute risk) that may be conducive to differential clinical decisions in people from different race groups. Under these circumstances, the estimates from the CVD risk calculator could be ‘flagged’ to alert the clinician about entering a zone of potential ‘race-based’ medicine. Examples of such ‘flags’ could be a zone for risk estimates where Black-white differences (or the converse) in absolute risk exceed 2.5% and/or the corresponding relative risks exceed 1.5, a magnitude generally regarded of major significance in epidemiology for most exposures.

Last, developing a causal framework for risk prediction entails estimating the risk for a person whose risk factor profile we would like to alter with an intervention of known effect size in a group of persons with similar characteristics; this requires counterfactual risk prediction, which could delineate the risk experienced by a similar person with an altered (post-intervention) risk profile. Such counterfactual prediction will clarify whether (and to what extent) we can mitigate the higher CVD risk experienced by Blacks by intervening on their risk profile. The PCE were developed using only observational (factual) data and did not include estimation of reduced risk after treatment (counterfactual data). Thus, the PCE could be improved by including longitudinal data where we take into account risk modification with interventions and by replacing ‘race’ with the underlying truly causal factors that can be intervened upon.

### Study Limitations

We studied numerous risk factor combinations determined by our choice of pragmatic cut points for ‘binning’ the continuous range of numerical risk factors. An *in silico* approach facilitated evaluation of PCE-based risk estimation across a very wide range of risk factor profiles, not readily feasible using data from individual studies or pooled cohorts. Yet, our *in silico* results may be questioned as theoretical. Accordingly, we repeated our analyses using a range of risk factors within the normal range. Furthermore, we evaluated two independent community-based samples spanning a wide age range to support our findings. Overall, our analysis of FHS and NHANES data confirm that differences in the 10-year absolute risk of CVD between Black and white people with identical profiles may occur frequently within the typical numerical range of risk factors in community-dwelling ambulatory individuals.

## Conclusions

The race term in the PCE can result in substantially divergent absolute and relative risks for CVD for Black vs. white persons with identical risk factor profiles. Such divergence in estimated CVD risk may introduce race-related variations in physician recommendations for CVD prevention.

## Data Availability

All data used in the manuscript are available at BiolINCC and NCHS website

https://biolincc.nhlbi.nih.gov/home/

https://www.n.cdc.gov/nchs/nhanes/continuousnhanes/default.aspx?BeginYear=2017

## Acknowledgments

This work is supported by Contracts NO1-HC-25195, HHSN268201500001I and 75N92019D00031 and U01HL146382 from the National Heart, Lung and Blood Institute. Dr. Vasan is supported in part by the Evans Medical Foundation and the Jay and Louis Coffman Endowment from the Department of Medicine, Boston University School of Medicine.

The funding agency (National Heart, Lung and Blood Institute) has no role in: the design and conduct of this report; the analysis, and interpretation of the data; preparation, review, or approval of the manuscript; and decision to submit the manuscript for publication.

