## Supplementary Appendix for "Differences in estimates for ten-year risk of cardiovascular disease in Black versus white persons with identical risk factor profiles using pooled cohort equations"

**Supplementary Appendix: Table of Contents**

| **Content** | **Page** |
| --- | --- |
| **Table S1.** Summary statistics for the calculated numerical risk factors for men and women (data for pooled races) | 2 |
| **Table S2.** Summary statistics for the calculated numerical risk factors when a difference in pooled cohort equation-based 10-year CVD risk is present in Black versus white men. | 3 |
| **Table S3.** Non-smoking Men without diabetes or hypertension treatment with divergent Black versus white pooled cohort equation-based 10-year CVD risk when their risk factor profile is identical and values of systolic blood pressure, total and high density lipoprotein cholesterol HDL are in normal range. | 4 |
| **Table S4.** Summary statistics for the calculated numerical risk factors when a difference in pooled cohort equation-based 10-year CVD risk is present in Black versus white women. | 5 |
| **Table S5.** Non-smoking women without diabetes or hypertension treatment with divergent Black versus white pooled cohort equation-based 10-year CVD risk when their risk factor profile is identical and values of systolic blood pressure, total and high density lipoprotein cholesterol HDL are in normal range. | 6 |
| **Table S6.** Median [Q1, Q3] for numerical risk factors and percentages for binary risk factors in participants of the FHS Generation 3 cohort at their first examination cycle | 7 |
| **Table S7.** Median [Q1, Q3] for numerical risk factors and percentages for binary risk factors in participants of the NHANES 2017-2018 cycle | 8 |
| **Figure S1.** Patterns in risk factors across four different groups of pooled cohort equation-based 10-year CVD risk estimates in men | 9-10 |
| **Figure S2.** Risk differences in men by level of risk factors (Left column Black men at risk and white men not at risk; Right column white men at risk and Black men not at risk). ‘At risk’ indicates pooled cohort equation-based 10-year CVD risk exceeds 7.5%. | 11-13 |
| **Figure S3.** Patterns in risk factors across four different groups of pooled cohort equation-based 10-year CVD risk estimates in women | 14-15 |
| **Figure S4.** Risk differences in women by level of risk factors (Left column Black women at risk and white women not at risk; Right column white women at risk and Black women not at risk). ‘At risk’ indicates pooled cohort equation-based 10-year CVD risk exceeds 7.5%. | 16-18 |

**Table S1.** **Summary statistics for the calculated numerical risk factors for men and women (data for pooled races)**

| **Risk Factor** | **Men** | | | | | **Women** | | | | |
| --- | --- | --- | --- | --- | --- | --- | --- | --- | --- | --- |
|  | **N^a^** | **Median** | **IQR** | **Minimum** | **Maximum** | **N^a^** | **Median** | **IQR** | **Minimum** | **Maximum** |
| Age, years | 30565 | 55 | 15 | 40 | 80 | 29515 | 55 | 15 | 40 | 80 |
| Total Cholesterol, mg/dl | 30565 | 210 | 80 | 130 | 290 | 29515 | 210 | 80 | 130 | 290 |
| HDL, mg/dl | 30565 | 60 | 40 | 20 | 90 | 29515 | 60 | 30 | 20 | 90 |
| Untreated SBP, mm Hg | 17898 | 140 | 50 | 100 | 200 | 16311 | 140 | 50 | 100 | 200 |
| Treated SBP, mm Hg | 12667 | 130 | 40 | 100 | 180 | 13204 | 130 | 40 | 100 | 180 |

^a^N= number of risk factor combinations. IQR= interquartile range. HDL= high-density lipoprotein cholesterol. SBP= systolic blood pressure.

**Table S2. Summary statistics for the calculated numerical risk factors when a difference in pooled cohort equation-based 10-year CVD risk is present in Black versus white men.**

| **Risk Factor** | **Black men at risk*,**  **but white men are not** | | | | | **White men at risk*,**  **but Black men are not** | | | | |
| --- | --- | --- | --- | --- | --- | --- | --- | --- | --- | --- |
|  | **N^a^** | **Median** | **IQR** | **Minimum** | **Maximum** | **N^a^** | **Median** | **IQR** | **Minimum** | **Maximum** |
| Age, years | 6357 | 45 | 10 | 40 | 65 | 391 | 50 | 15 | 40 | 75 |
| Total Cholesterol, mg/dl | 6357 | 190 | 80 | 130 | 290 | 391 | 250 | 60 | 130 | 290 |
| HDL, mg/dl | 6357 | 70 | 30 | 20 | 90 | 391 | 30 | 30 | 20 | 90 |
| Untreated SBP, mm Hg | 2667 | 160 | 50 | 100 | 200 | 375 | 110 | 20 | 100 | 180 |
| Treated SBP, mm Hg | 3690 | 140 | 40 | 100 | 180 | 16 | 100 | 10 | 100 | 130 |

^a^N= number of risk factor combinations. IQR= interquartile range. HDL= high-density lipoprotein cholesterol. SBP= systolic blood pressure.

*‘at risk’ indicates 10-year CVD risk exceeds 7.5%.

.

**Table S3. Non-smoking Men without diabetes or hypertension treatment with divergent Black versus white pooled cohort equation-based** **10-year CVD risk when their risk factor profile is identical and values of systolic blood pressure, total and high density lipoprotein cholesterol HDL are in normal range.**

| **Age, years** | **Total Cholesterol, mg/dl** | **HDL, mg/dl** | **SBP, mm Hg** | **10-year estimated CVD risk, white men** | **10-year estimated CVD risk, Black men** | **Absolute Risk Difference** | **Relative Risk** |
| --- | --- | --- | --- | --- | --- | --- | --- |
| 60 | 170 | 40 | 120 | 7.8183 | 7.46116 | 0.35714 | 1.04787 |
| 65 | 130 | 40 | 110 | 8.3231 | 7.17388 | 1.14924 | 1.16020 |
| 65 | 130 | 60 | 120 | 7.6200 | 7.40490 | 0.21512 | 1.02905 |
| 65 | 150 | 40 | 100 | 7.8330 | 6.33257 | 1.50047 | 1.23694 |
| 65 | 150 | 40 | 110 | 9.1992 | 7.47856 | 1.72068 | 1.23008 |
| 65 | 150 | 50 | 110 | 8.0850 | 7.00116 | 1.08385 | 1.15481 |
| 65 | 150 | 70 | 120 | 7.7045 | 7.37570 | 0.32877 | 1.04458 |
| 65 | 170 | 40 | 100 | 8.5520 | 6.56833 | 1.98366 | 1.30200 |
| 65 | 170 | 50 | 100 | 7.5129 | 6.14709 | 1.36579 | 1.22219 |
| 65 | 170 | 50 | 110 | 8.8260 | 7.26083 | 1.56513 | 1.21556 |
| 65 | 170 | 60 | 110 | 7.9412 | 6.87952 | 1.06164 | 1.15432 |
| 70 | 130 | 40 | 100 | 10.7524 | 7.24719 | 3.50523 | 1.48367 |
| 70 | 130 | 50 | 100 | 9.7267 | 6.78401 | 2.94273 | 1.43377 |
| 70 | 130 | 60 | 100 | 8.9581 | 6.42686 | 2.53120 | 1.39385 |
| 70 | 130 | 70 | 100 | 8.3535 | 6.13911 | 2.21440 | 1.36070 |
| 70 | 130 | 70 | 110 | 9.8056 | 7.25147 | 2.55410 | 1.35222 |
| 70 | 150 | 50 | 100 | 10.4588 | 7.07277 | 3.38599 | 1.47874 |
| 70 | 150 | 60 | 100 | 9.6354 | 6.70097 | 2.93440 | 1.43791 |
| 70 | 150 | 70 | 100 | 8.9874 | 6.40138 | 2.58603 | 1.40398 |
| 70 | 170 | 50 | 100 | 11.1412 | 7.33499 | 3.80622 | 1.51891 |
| 70 | 170 | 60 | 100 | 10.2672 | 6.94993 | 3.31730 | 1.47731 |
| 70 | 170 | 70 | 100 | 9.5791 | 6.63960 | 2.93946 | 1.44272 |
| 75 | 130 | 70 | 100 | 13.0941 | 7.23701 | 5.85707 | 1.80932 |

Normal range: for systolic blood pressure, 100-130 mm Hg; total cholesterol = 130-170 mg/dl; HDL cholesterol, 40-70 mg/dl.

SBP= systolic blood pressure. HDL= high-density lipoprotein cholesterol.

**Table S4. Summary statistics for the calculated numerical risk factors when a difference in pooled cohort equation-based** **10-year CVD risk is present in Black versus white women.**

| **Risk Factor** | **Black women at risk*,**  **but white women are not** | | | | | **White women at risk*,**  **but Black women are not** | | | | |
| --- | --- | --- | --- | --- | --- | --- | --- | --- | --- | --- |
|  | **N^a^** | **Median** | **IQR** | **Minimum** | **Maximum** | **N^a^** | **Median** | **IQR** | **Minimum** | **Maximum** |
| Age, years | 6543 | 50 | 10 | 40 | 70 | 318 | 45 | 30 | 40 | 80 |
| Total Cholesterol, mg/dl | 6543 | 190 | 100 | 130 | 290 | 318 | 230 | 100 | 130 | 290 |
| HDL, mg/dl | 6543 | 60 | 40 | 20 | 90 | 318 | 30 | 20 | 20 | 90 |
| Untreated SBP, mm Hg | 3542 | 160 | 40 | 100 | 200 | 186 | 110 | 20 | 100 | 170 |
| Treated SBP, mm Hg | 3001 | 150 | 40 | 100 | 180 | 132 | 110 | 10 | 100 | 140 |

^a^N= number of risk factor combinations. IQR= interquartile range. HDL= high-density lipoprotein cholesterol. SBP= systolic blood pressure.

*‘at risk’ indicates 10-year CVD risk exceeds 7.5%.

**Table S5. Non-smoking women without diabetes or hypertension treatment with divergent Black versus white pooled cohort equation-based** **10-year CVD risk when their risk factor profile is identical and values of systolic blood pressure, total and high density lipoprotein cholesterol HDL are in normal range.**

| **Age, years** | **Total Cholesterol, mg/dl** | **HDL, mg/dl** | **SBP, mm Hg** | **10-year estimated CVD risk, white women** | **10-year estimated CVD risk, Black women** | **Absolute Difference** | **Relative Risk** |
| --- | --- | --- | --- | --- | --- | --- | --- |
| 70 | 150 | 70 | 120 | 7.37919 | 8.19782 | 0.81863 | 1.11094 |
| 70 | 150 | 80 | 120 | 7.19237 | 8.29485 | 1.10247 | 1.15328 |
| 70 | 170 | 50 | 110 | 6.93574 | 7.56446 | 0.62872 | 1.09065 |
| 70 | 170 | 60 | 110 | 6.69650 | 7.68738 | 0.99088 | 1.14797 |
| 70 | 170 | 70 | 110 | 6.50045 | 7.79281 | 1.29236 | 1.19881 |
| 70 | 170 | 80 | 110 | 6.33511 | 7.88525 | 1.55014 | 1.24469 |
| 70 | 170 | 80 | 120 | 7.46620 | 9.28096 | 1.81476 | 1.24306 |
| 70 | 130 | 40 | 120 | 7.87177 | 6.85531 | 1.01645 | 1.14827 |
| 70 | 130 | 50 | 120 | 7.54231 | 6.99244 | 0.54988 | 1.07864 |
| 75 | 130 | 40 | 100 | 9.79861 | 7.04589 | 2.75272 | 1.39068 |

Normal range: for systolic blood pressure, 100-130 mm Hg; total cholesterol = 130-170 mg/dl; HDL cholesterol, 50-80 mg/dl. HDL= high-density lipoprotein cholesterol.

**Table S6. Median [Q1, Q3] for numerical risk factors and percentages for binary risk factors in participants of the FHS Generation 3 cohort at their first examination cycle**

| **Risk factors** | **Overall**  **(N=4086)** | **Men**  **(N=1907)** | **Women (N=2179)** |
| --- | --- | --- | --- |
| Age, years | 40 [34, 46] | 40 [34, 47] | 40 [34, 46] |
| Total cholesterol, mg/dl | 186 [165, 210] | 191 [168, 216] | 182 [162, 205] |
| High-density lipoprotein cholesterol, mg/dl | 52 [43, 64] | 45 [38, 54] | 60 [50, 70] |
| Systolic blood pressure, mm Hg | 115 [107, 124] | 119 [112, 128] | 111 [103, 121] |
| Hypertension treatment, % | 8.44 | 9.81 | 7.23 |
| Smoking, % | 15.1 | 15.9 | 14.3 |
| Diabetes mellitus, % | 2.99 | 3.78 | 2.29 |

Q1, Q3 indicate first and third quartile cutpoints.

**Table S7. Median [Q1, Q3] for numerical risk factors and percentages for binary risk factors in participants of the NHANES 2017-2018 cycle**

| **Risk factors** | **Overall**  **(N=1065)** | **Men**  **(N=611)** | **Women  (N=454)** |
| --- | --- | --- | --- |
| Age, years | 61 [46, 71] | 63 [49, 72] | 57 [42, 68] |
| Total cholesterol, mg/dl | 184 [158, 214] | 177 [148, 208] | 193 [169, 221] |
| High-density lipoprotein cholesterol, mg/dl | 50 [41, 60] | 46 [39, 56] | 54 [46, 66] |
| Systolic blood pressure, mm Hg | 127 [116, 141] | 127 [117, 141] | 125 [114, 141] |
| Hypertension treatment, % | 33.1% | 35.8% | 29.3% |
| Smoking, % | 39.0% | 34.5% | 44.9% |
| Diabetes mellitus, % | 20.0% | 22.4% | 16.7% |
| Black Race, % | 33.6% | 33.2% | 34.1% |

Q1, Q3 indicate first and third quartile cutpoints.

**Figure S1.** Patterns in risk factors across four different groups of pooled cohort equation-based 10-year CVD risk estimates in men

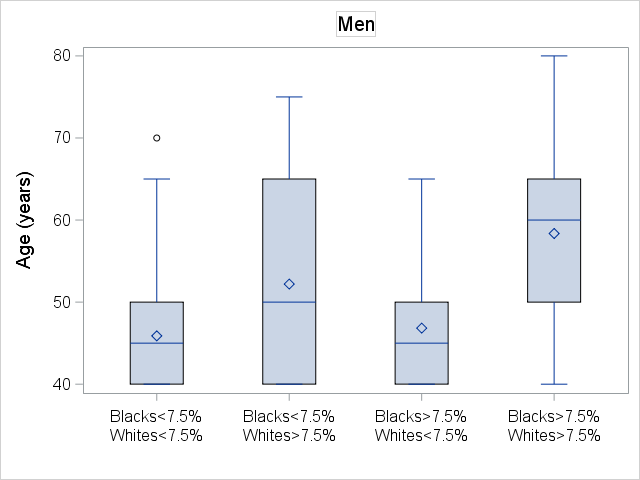

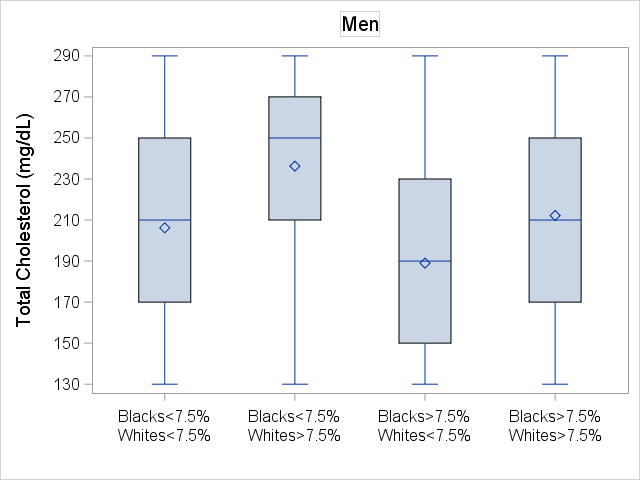

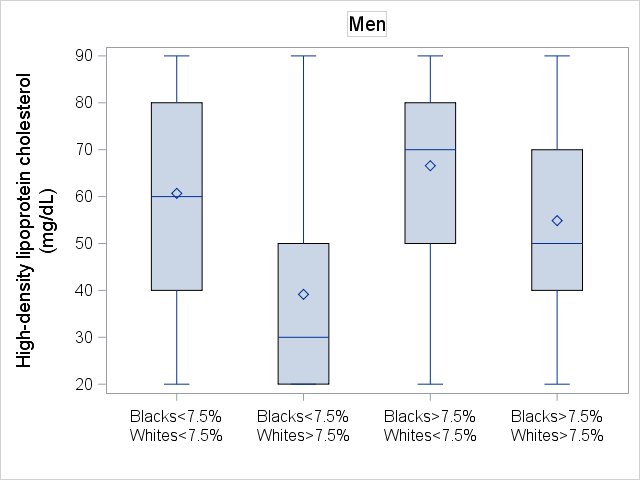

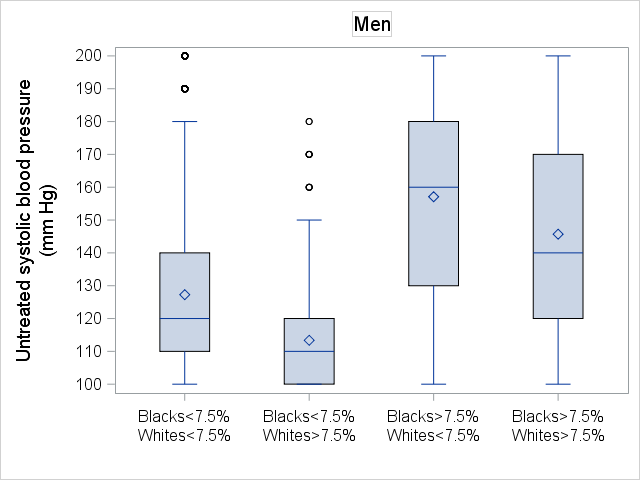

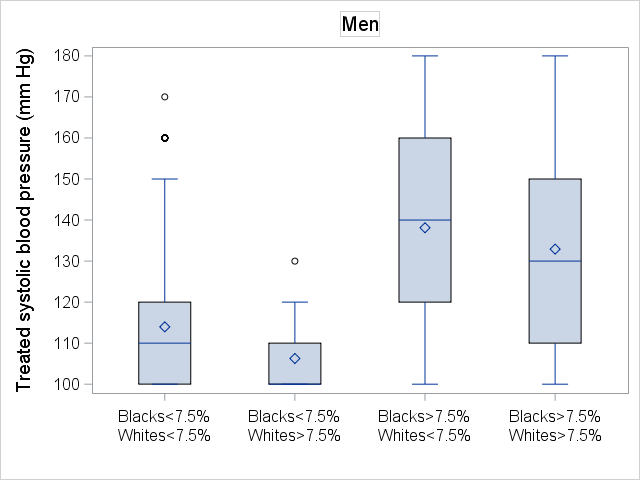

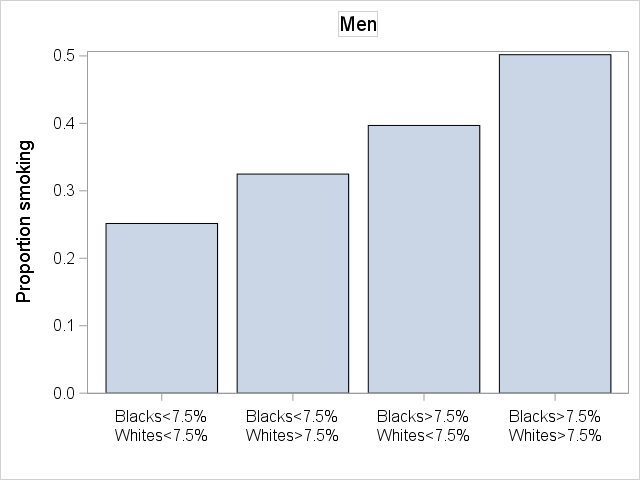

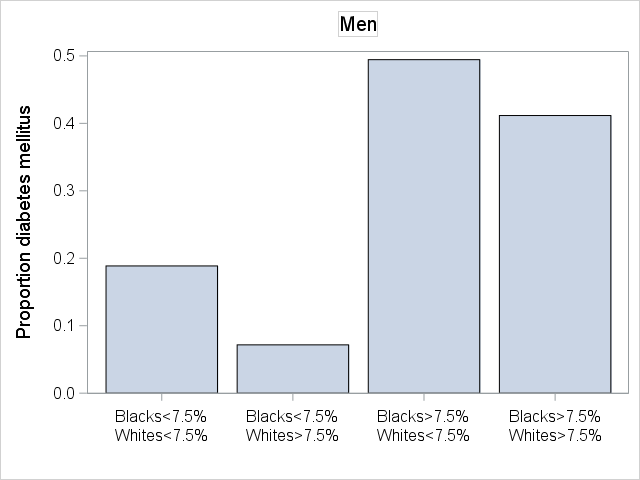

**Figure S2.** Risk differences in men by level of risk factors (Left column Black men at risk and white men not at risk; Right column white men at risk and Black men not at risk). ‘At risk’ indicates pooled cohort equation-based 10-year CVD risk exceeds 7.5%.

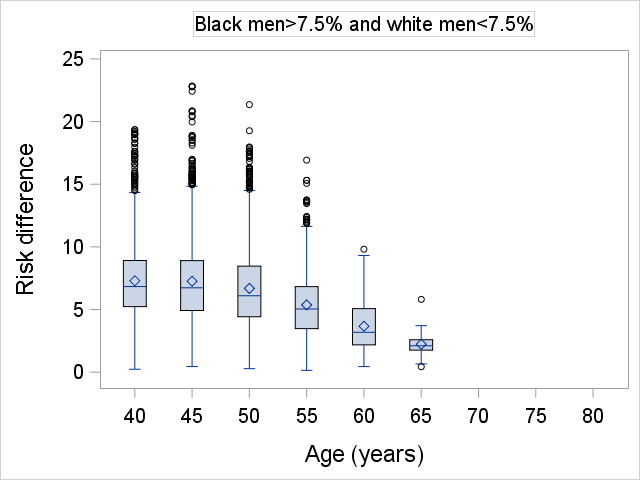

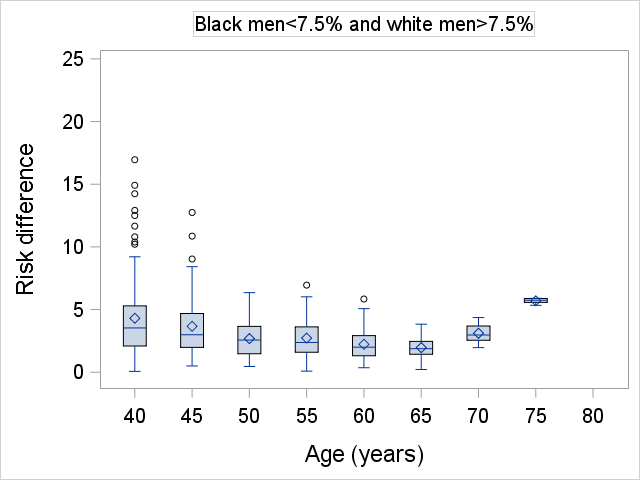

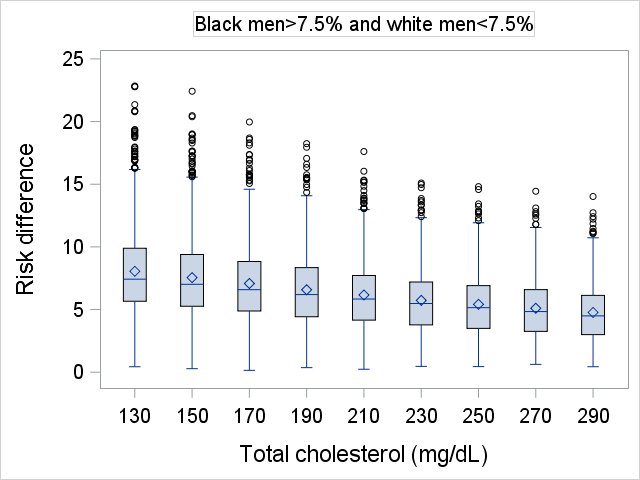

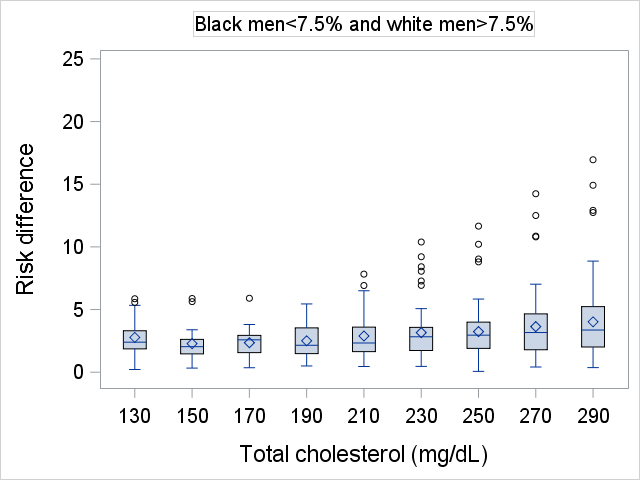

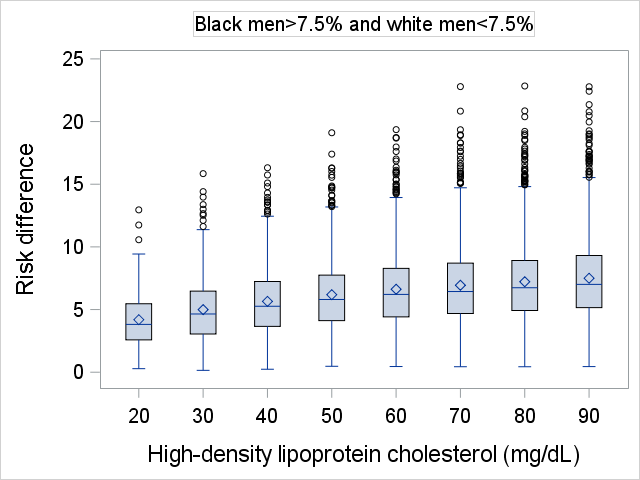

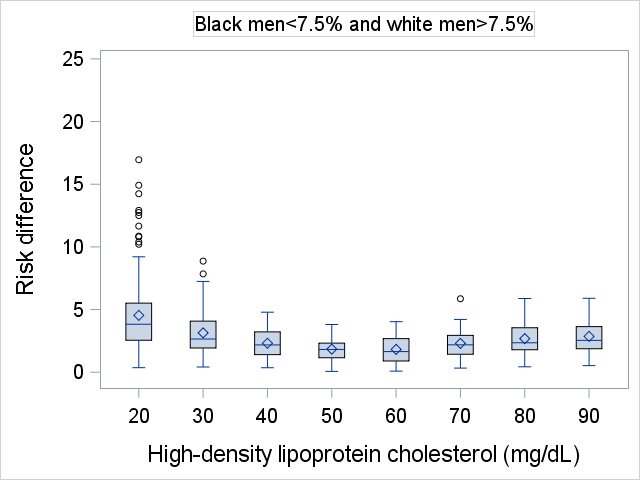

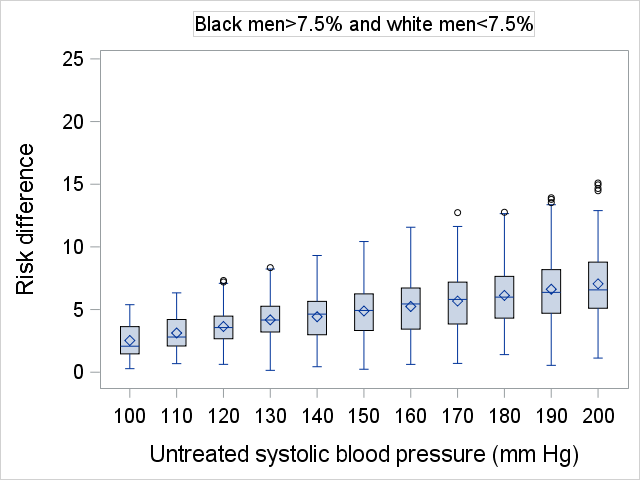

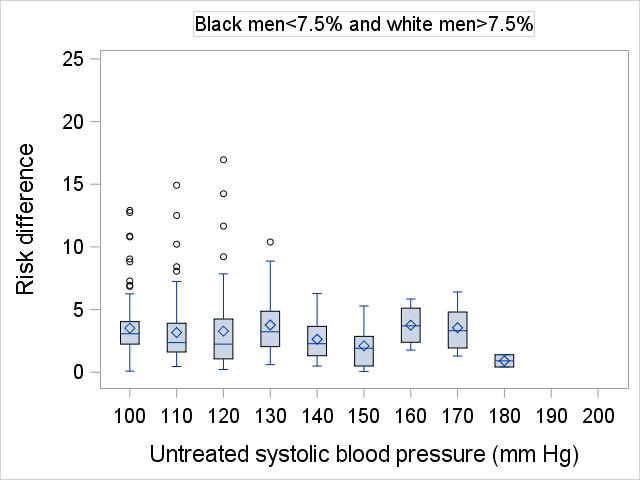

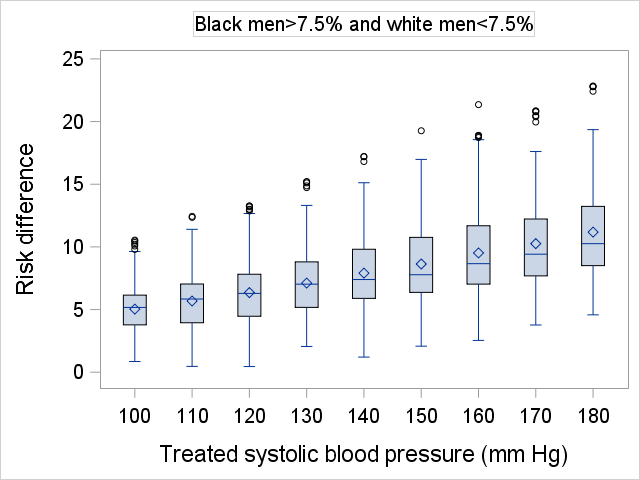

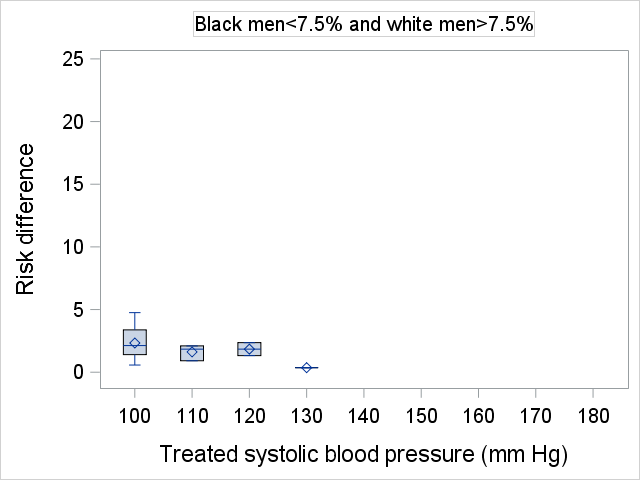

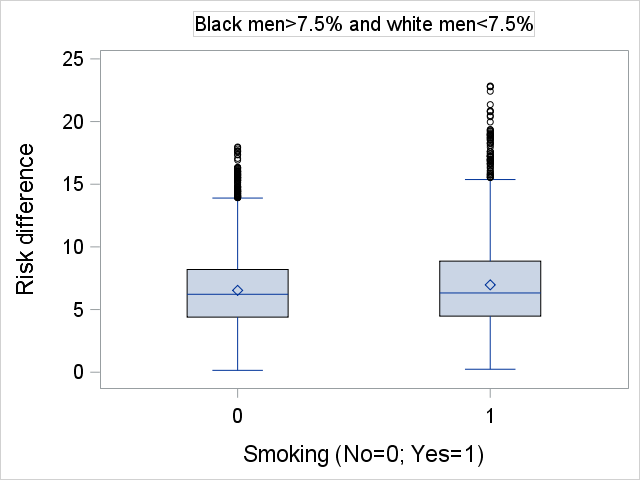

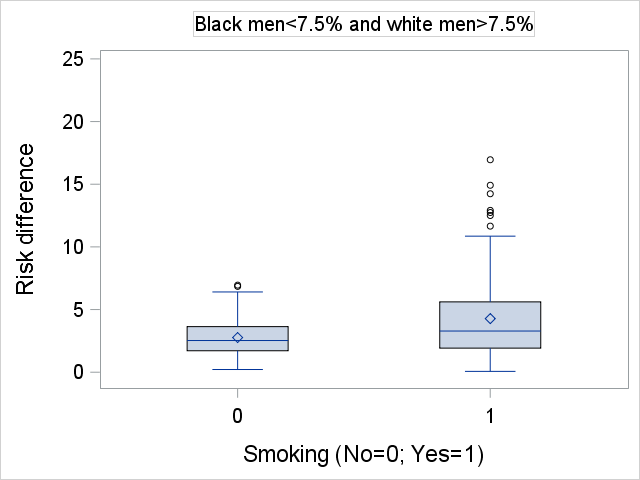

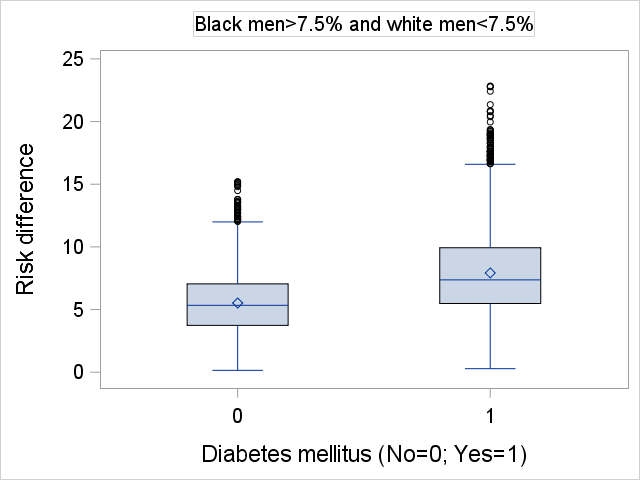

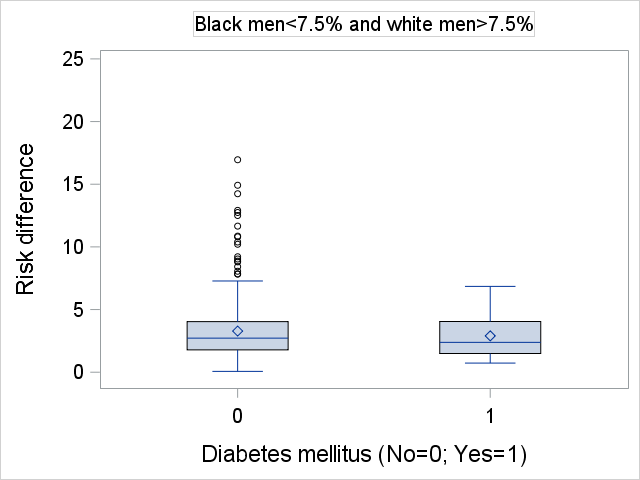

**Figure S3.** Patterns in risk factors across four different groups of pooled cohort equation-based 10-year CVD risk estimates in women

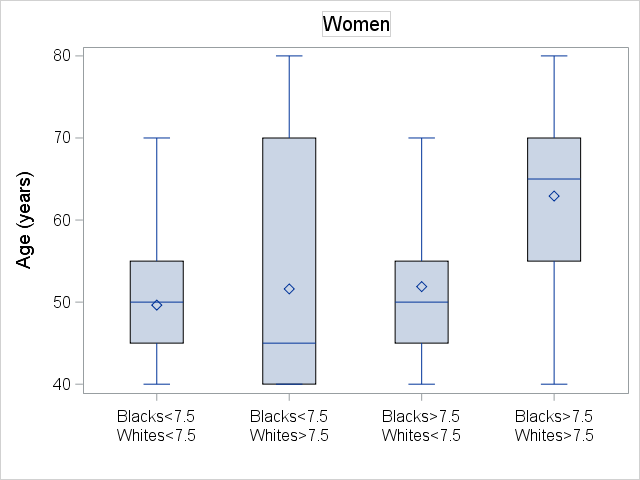

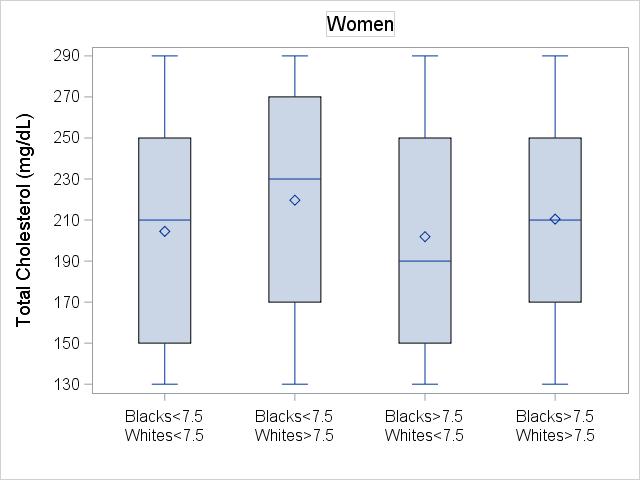

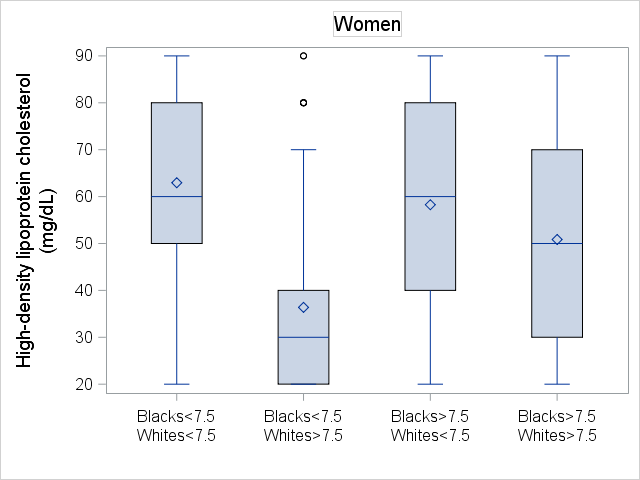

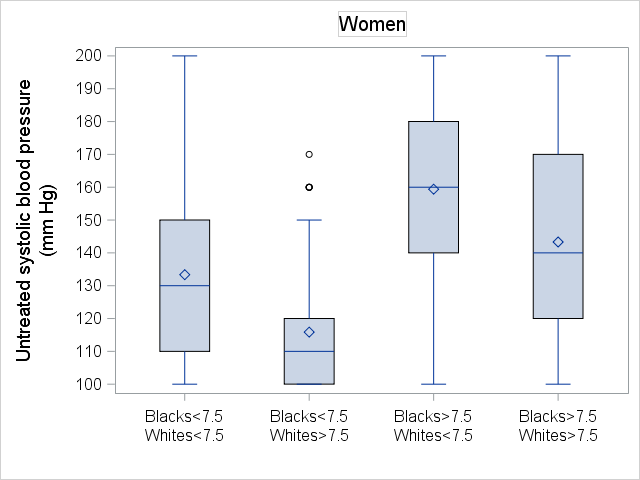

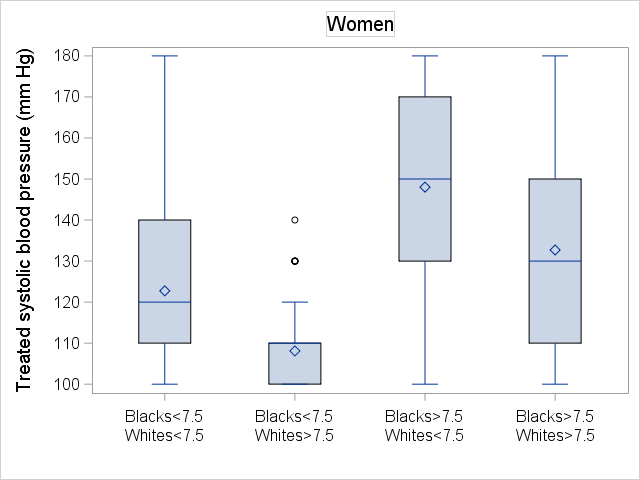

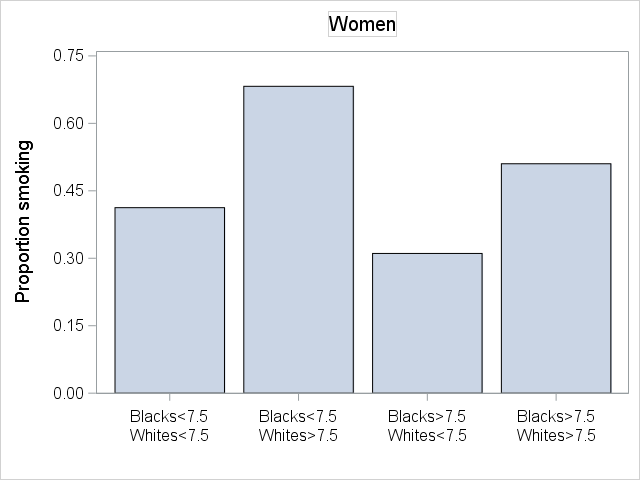

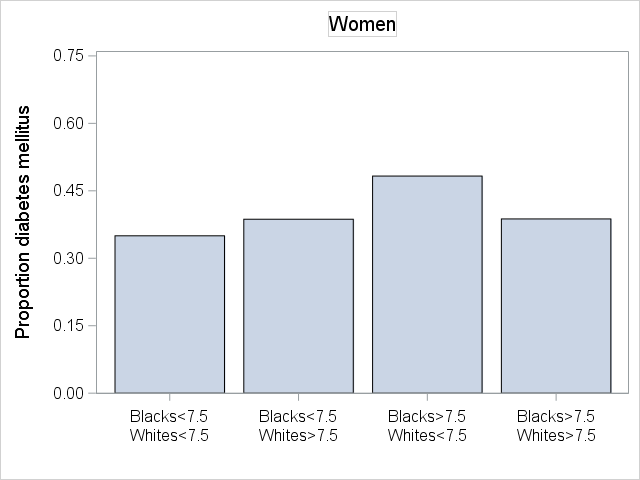

**Figure S4.** Risk differences in women by level of risk factors (Left column Black women at risk and white women not at risk; Right column white women at risk and Black women not at risk). ‘At risk’ indicates pooled cohort equation-based 10-year CVD risk exceeds 7.5%.

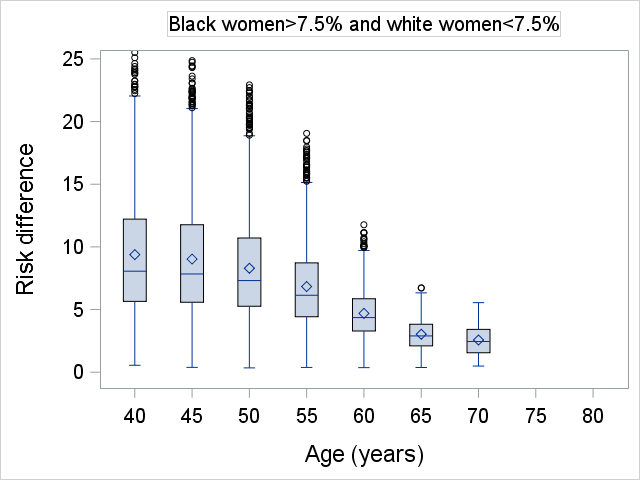

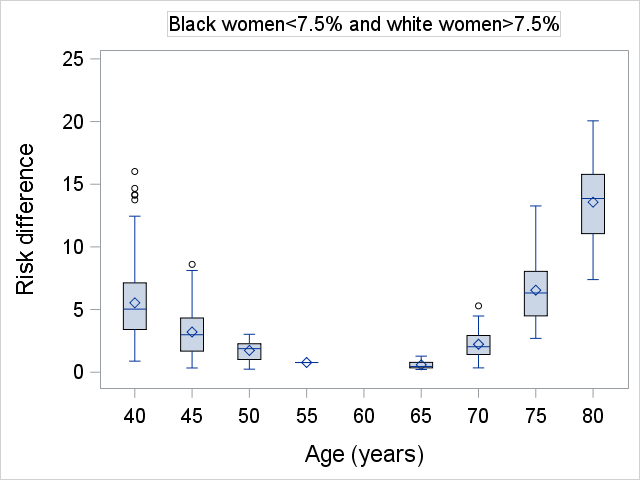

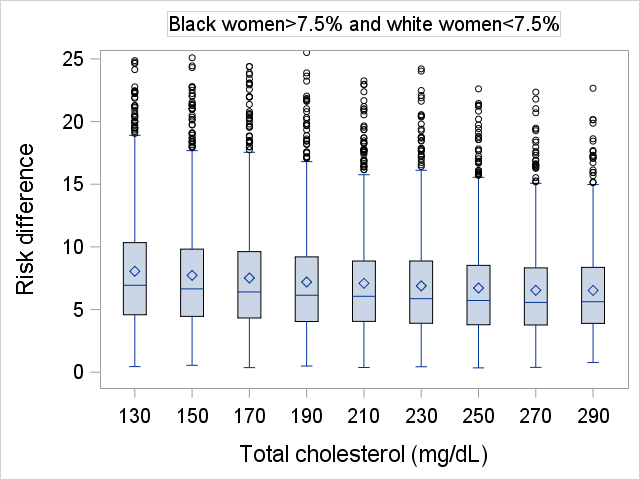

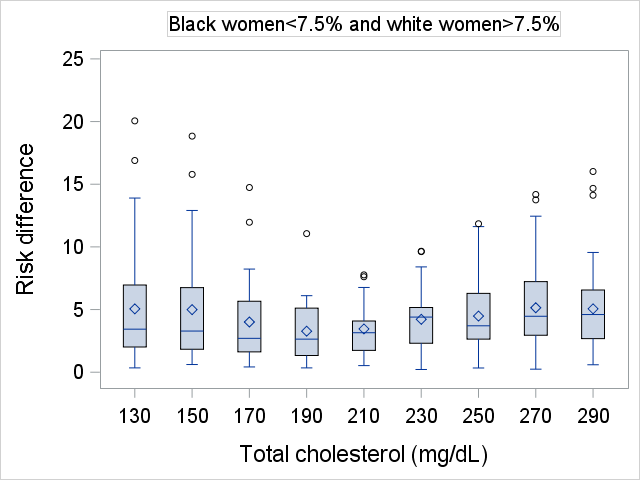

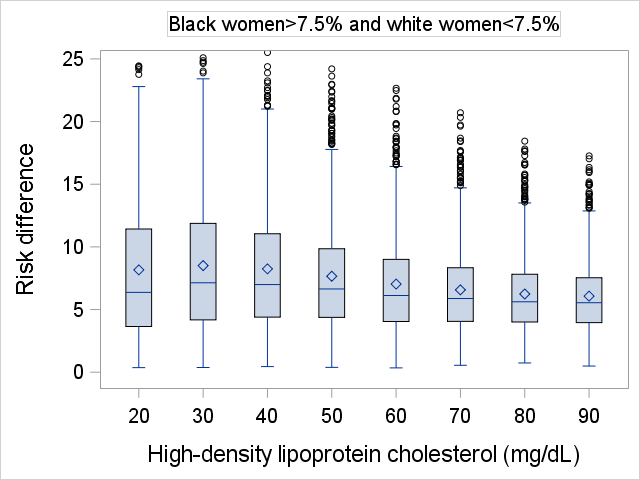

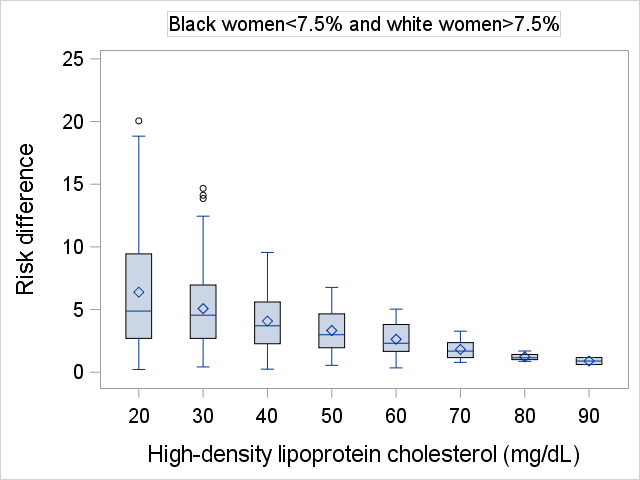

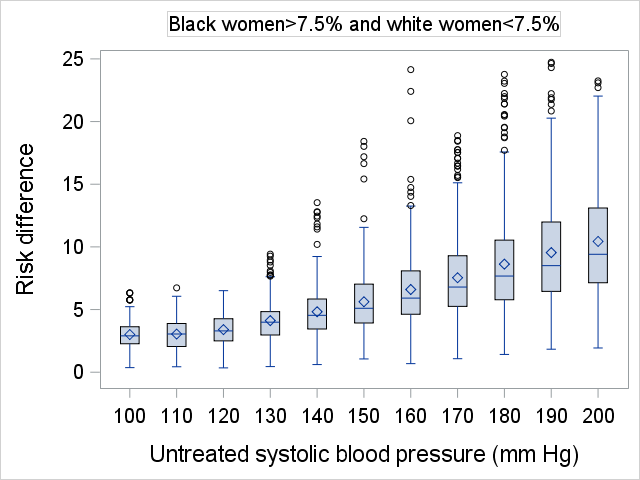

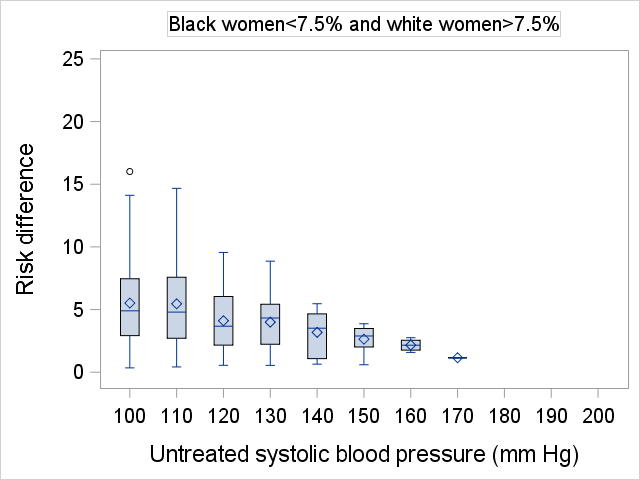

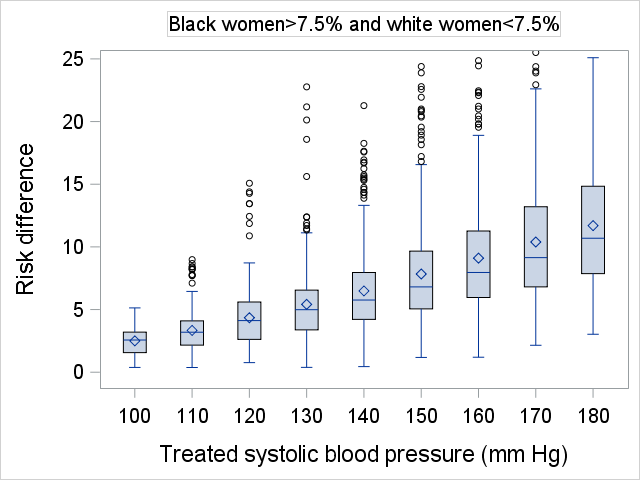

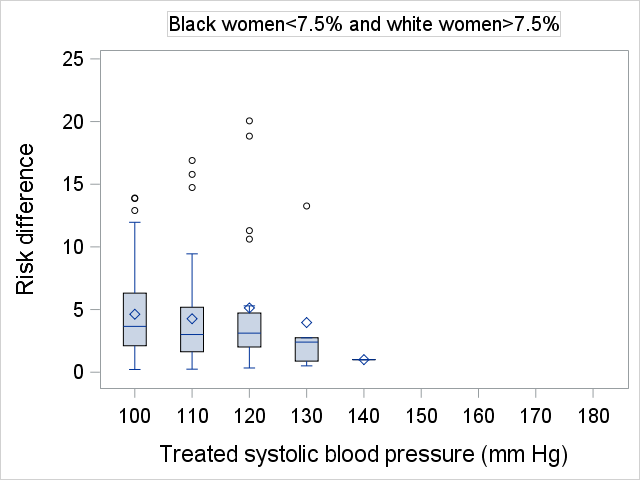

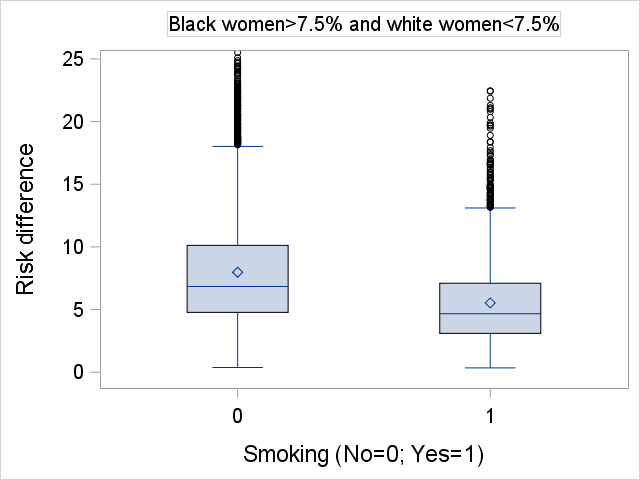

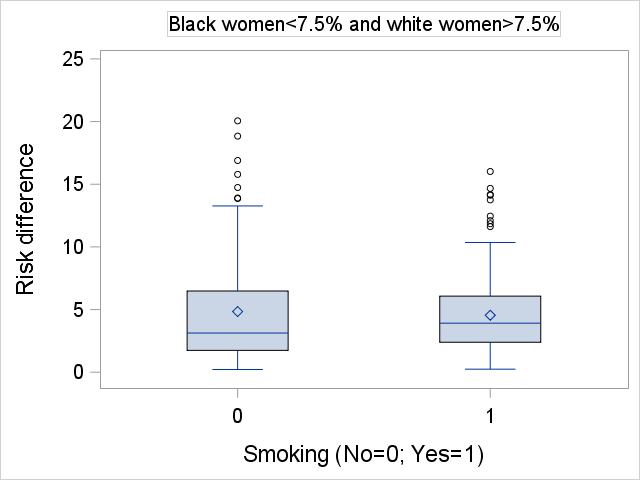

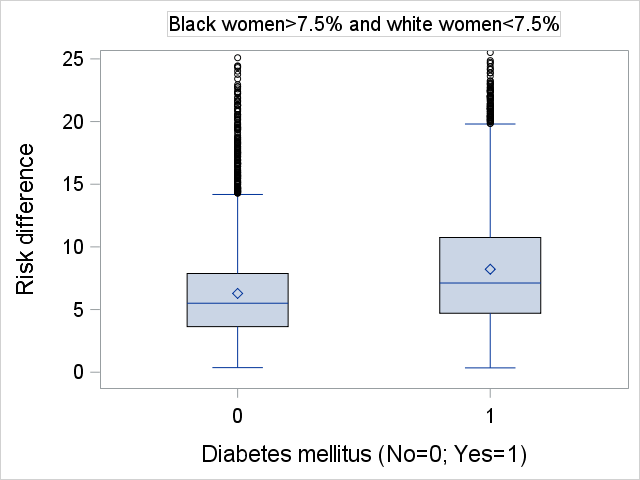

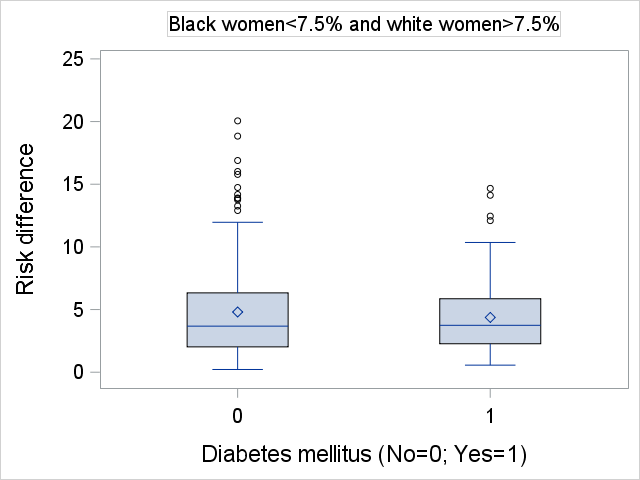
